# Much more than a name change: Impact of the new steatotic liver disease nomenclature on clinical algorithms and disease classification in U.S. adults and adolescents

**DOI:** 10.1101/2023.08.04.23293664

**Authors:** Ning Ma, Meena Bansal, Jaime Chu, Andrea D. Branch

## Abstract

**Background and Aims:** The newly proposed nomenclature for steatotic liver diseases (SLD) aims to reduce the stigma associated with “non-alcoholic fatty liver disease” (NAFLD), increase awareness, and provide a framework for delineating pathogenic pathways.

**Approach and Results:** We projected the new nomenclature’s diagnostic scheme onto National Health and Nutrition Examination Survey (NHANES) data and determined SLD prevalence, fibrosis risk factors, subtypes, and consistency with previous classifications. Steatosis grade and fibrosis stage were estimated from vibration controlled transient elastography (VCTE). At a threshold of 240 dB/m, 62.1% [95% confidence interval (CI), 59.8-64.3%] of adults (≥ 20 years) and 30.5% (95% CI, 27.1-34.0%) of adolescents (12-19 years) had SLD. By American Gastroenterological Association criteria, 19.3 million (95% CI, 15.8-22.8) adults with SLD qualify for hepatology referral. Over 98% of adults but only 85% of adolescents with NAFLD met criteria for definite MASLD. Significant fibrosis (≥ 8.6 kPa) occurred in 13.5 million (95% CI, 10.9-16.2) adults with MASLD; risk factors varied by race and ethnicity. Significant fibrosis occurred in over 1.5 million adults without any identified LD and was associated with lead (Pb) exposure, odds ratio = 3.89 (95% CI, 2.00-7.56).

**Conclusions:** The overarching term, SLD, changes the diagnostic algorithm and creates an umbrella classification that highlights the extraordinary prevalence of liver steatosis. The more precise nomenclature establishes a valuable patient-centric platform for research and clinical care, clarifying risk groups and risk factors, including adolescents with NAFLD but without definite MASLD and adults without SLD in whom toxic exposures may increase fibrosis risk.

## Introduction

Recently, members of multinational liver societies and patient advocacy groups used a modified Delphi approach to evaluate steatotic liver disease (SLD) nomenclature^1^. The expert panel aimed to reduce the stigma associated with the words “non-alcoholic” and “fatty” in the term non-alcoholic fatty liver disease (NAFLD), while also increasing awareness of metabolic dysfunction as a driver of LD, stimulating research, and supporting the uninterrupted development of pharmaceuticals and biomarkers. The panel issued a statement reporting on its findings and proposing a new diagnostic scheme in which liver steatosis and cardiometabolic risk factors (CMRF) are key determinants^1, 2^. In the new algorithm, patients are first divided into those with and without SLD. Patients with SLD are then divided into those with and without at least one of the designated CMRFs.

Patients with SLD and at least one CMRF who consume no more than minimal alcohol [up to 140 g/week (women), 210 g/week (men)] have metabolic dysfunction-associated liver disease (MASLD) if they have no other identifiable cause of LD. MASLD is the proposed replacement term for NAFLD. As reported by the nomenclature panel^1^, in the Liver Investigation: Testing Marker Utility in Steatohepatitis (LITMUS) study, nearly all the patients with NAFLD in this international cohort have at least one of the five designated CMRFs and thus meet the diagnostic criteria of MASLD, but to our knowledge, the concordance of NAFLD and MASLD has not previously been examined in adults and adolescents in the United States.

The new diagnostic algorithm divides many liver diseases, including viral hepatitis, into multiple categories, e.g., viral hepatitis ± steatosis and ± CMRF. This is expected to increase awareness of steatosis and insulin resistance as disease accelerants across all liver disease etiologies. Additionally, by segregating patients with SLD but no CMRFs into a separate subgroup, the new diagnostic scheme can be expected to advance research into the risk factors for and the natural history of cryptogenic liver disease. Alcohol-related liver diseases are classified differently than in the past. Patients who meet all criteria for MASLD except that they consume a moderately higher amount of alcohol [140-350 g/week (women), 210-420 g/week (men)] have a new more precise diagnosis, metabolic dysfunction-associated alcohol-associated liver disease (MetALD). This new designation emphasizes that the contributions of metabolic dysfunction and alcohol toxicity need to be assessed in each individual patient, as their relative importance varies along a spectrum.

This study was undertaken to project the new nomenclature’s diagnostic scheme onto the population of the United States using data available in the National Health and Nutrition Examination Survey (NHANES). Major advantages of this dataset include the high-quality and completeness of the data, the over-sampling of non-Hispanic Black (NHB), Mexican American (MA), and other (O) racial and ethnic groups, and the availability of statistical methods that allow findings to be extrapolated to the housed civilian population of the entire United States^3^. The nomenclature panel did not define what “constitutes hepatic steatosis”, but noted that “in reality most, if not all, patients will usually have imaging at some point.”^1^ Given the pre-eminent role non-invasive determination of liver steatosis is expected to play, the cost-effectiveness of the controlled attenuation parameter (CAP) of vibration controlled transient elastography (VCTE), the greater sensitivity of VCTE-CAP compared to ultrasound for the detection of increased intrahepatic lipids, and the availability of VCTE data in NHANES, we used VCTE-CAP to estimate hepatitis steatosis grade.

Applying a 240 dB/m threshold for steatosis grade 1, as recommended by the major supplier (EchoSens)^4^, about 30% of adolescents and 62% of adults had SLD. Among people with SLD and NAFLD, over 98% of adults, but only 85% of adolescents had definite MASLD. Applying 8.6 kPa as the threshold for fibrosis stage F2 and higher^5^, almost 90% of people with significant fibrosis were in the SLD/CMRF group. Importantly, associations between CMRFs and significant liver fibrosis varied by race/ethnicity. In the No-SLD group, most people with significant fibrosis were in the subgroup that lacks any defined cause of liver disease. Our results demonstrate the value of the overarching term SLD and the new diagnostic algorithm that accompanies it, as they focus attention on steatosis and metabolic dysfunction as important risk factors. Our findings establish that the more precise nomenclature provides a patient-centric framework for understanding liver disease pathogenesis and illustrates its utility for public health and workforce planning.

## Methods

### Study population and data sources

Data collection in NHANES follows standardized procedures approved by the National Center for Health Statistics Research Ethnic Review Board. IRB review was waived for analysis of de-identified NHANES data.^3^ NHANES 2017-March 2020 datasets with VCTE data and the NHANES III (1988-1994) dataset with abdominal ultrasound data were used; the public-use linked mortality file was obtained through 2019.^6^

### Indicators of SLD and significant fibrosis

SLD was defined by CAP ≥ 240 dB/m and additional threshold values. Significant fibrosis was defined by liver stiffness ≥ 8.6 kPa in adults and ≥ 7.4 kPa in adolescents^5, 7^. In sensitivity analysis, SLD was defined by ultrasound and classified as mild, moderate, or severe.

### Cardiometabolic risk factors (CMRFs) defined by the nomenclature review panel

Adults were evaluated for the CMRFs identified by the nomenclature review panel^1^: 1) Elevated adiposity, indicated by body mass index (BMI) ≥ 25 Kg/m^2^ or waist circumference (WC) > 80 cm (women) and > 94 cm (men); 2) Prediabetes/diabetes, indicated by fasting plasma glucose (FPG) ≥ 100 mg/dL or hemoglobin A1c (HbA1c) ≥ 5.7% or self-reported diagnosis of diabetes and treatment; 3) Elevated blood pressure, indicated by systolic blood pressure (SBP) ≥ 130 mmHg or diastolic blood pressure (DBP) ≥ 85 mmHg or use of anti-hypertensive medication; 4) Elevated triglycerides, indicated by plasma triglycerides ≥ 150 mg/dL or self-reported lipid lowering treatment; 5) Reduced plasma high-density lipoprotein cholesterol (HDL), indicated by plasma HDL ≤ 50 mg/dL (women) and ≤ 40 mg/dL (men) or self-reported lipid lowering treatment.

Adolescents (12-19 years of age) were evaluated for the CMRFs^1^ identified by the review panel:1) Elevated adiposity, indicated by BMI ≥ 85^th^ percentile for age/sex [BMI z score ≥ +1] or WC > 95^th^ percentile; 2) Prediabetes/diabetes, indicated by FPG ≥ 100 mg/dL or HbA1c ≥ 5.7% or self-reported diagnosis of diabetes and treatment; 3) Elevated blood pressure, indicated by BP ≥ 95^th^ percentile (12 ≤ age <13 years) and BP ≥ 130/85 mmHg (age ≥ 13 years) or use of anti-hypertensive medication; 4) Elevated triglycerides, indicated by plasma triglycerides ≥ 150 mg/dL or self-reported lipid lowering treatment; 5) Reduced HDL, indicated by plasma HDL ≤ 40 mg/dL or self-reported lipid lowering treatment.

### Liver disease definitions

NAFLD was SLD with no more than 140 g/week alcohol consumption for women and 210 g/week for men and no other discernable cause of liver disease. MASLD was SLD with at least one CMRF and no more than 140 g/week of alcohol consumption for women and 210 g/week for men and no other discernible cause. “Increased alcohol intake” was 140-350 g/week of alcohol for women and 210-420 g/week of alcohol for men over the past 12 months. MetALD was MASLD with increased alcohol intake. Alcohol associated liver disease (ALD) was > 350 g/week of alcohol for women or > 420 g/week for men over the past 12 months. Viral hepatitis (VH) was past/current infection with hepatitis B virus (HBV), positive core antibody or surface antigen; or hepatitis C virus (HCV), RNA or antibody. Cryptogenic SLD was SLD without a CMRF or any other identifiable cause. No Etiology Identified (NEI) was no discernable cause of liver disease and no SLD.

### Demographic variables

Self-reported sex (male/female) and race/ethincity [NHW, NHB, Mexican American (MA) and other (O) race (non-MA Hispanics and others)] were included. In NHANES 2017-March 2020 with VCTE data, the adult age range was 20 to over 80 years (people older than 80 were coded as 80 years); the adolscent age range was 12 to 19 years. In NHANES III, the age range of participants with ultrasound data was 20-74 years.

### Risk factors for significant fibrosis (stage F2 and greater)

Fibrosis risk factors were defined as in our previous study^8^. Briefly, diabetes was HbA1c ≥ 6.5%, and/or FPG ≥ 126 mg/dL and/or self-reported. Hypertension was SBP ≥ 130 mmHg, and/or DBP ≥ 80 mmHg, and/or use of anti-hypertensive medication. Past/current smokers answered “Yes” to the question “Have you smoked at least 100 cigarettes in your lifetime?”, never-smokers answered “No”. The responses to questions about alcohol consumption were used to create three mutually exclusive groups, lifetime abstainers (answered “No” for the question of “Ever had a drink of any kind of alcohol), former drinkers (had drinks in their lifetime but none in the past year), current drinkers (had drinks in the past year). Blood levels of lead (Pb) and cadmium (Cd) were analyzed as binary variables [quartiles (Q)1-3 versus Q4]. Poverty was defined as a family poverty-income-ratio below 1.0.

### Liver disease awareness

Liver disease awareness among adults was defined as follows: Participants answering “Yes” to the question “Has a doctor or other health professional ever told you had any liver condition”, were considered “aware”; those answering “No” or “Don’t know” were considered “unaware”.

### Statistical Analysis

All the analyses were performed following the guidelines established by NHANES^3^. The data were appropriately adjusted to account for the complex NHANES design by using survey commands in SAS OnDemand for Academics (SAS Institute Inc., Cary, NC, USA). The age standardization estimates were derived using the direct method and standardized to the 2000 US census population. Differences between groups were tested by univariate t statistics^9^. In order to determine the population counts in each etiology group, the weighted prevalence was calculated and subsequently multiplied by the estimated population in the United States; importantly, this weighted prevalence analysis differs slightly from the age-standardized-weighted prevalence described immediately above^3^. The population estimates were obtained from the American Community Survey^10^. Univariable and multivariable survey logistic regression with appropriate sample weights were used to examine the risk factors of significant fibrosis.

Survey-weighted adjusted univariable and multivariable cox proportional models were used to investigate the association between SLD subgroups and all-cause mortality. Variables with P value < 0.1 in the univariable analysis were included in multivariable models. Missing values less than 10% were addressed using multivariable imputation by chained equations^11^. Combined estimates using ten imputed datasets were calculated. Statistical significance was a two-sided P value <0.05.

## Results

### Projecting the new nomenclature’s diagnostic scheme onto the adult and adolescent populations of the United States

The primary analyses in this study were carried out on NHANES participants who had complete data for VCTE, CMRF, viral hepatitis, and alcohol consumption (for people more than 18 years of age). Separate analyses were carried out on adults (20 years and older) and adolescents (12-19 years). Sensitivity analysis was carried out on NHANES III participants 20 to 74 years of age with abdominal ultrasound data.

Flow diagrams show the diagnostic scheme of the new nomenclature applied to adults (Figure 1A) and adolescents (Figure 2A). The first branch point divides the population into people with and without SLD. By introducing the overarching term, SLD, and making it the first decision point in the diagnostic tree, the new nomenclature calls attention to the importance of liver steatosis at the personal and population levels and establishes a framework for investigating the natural history of liver disease in people with and without SLD.

**Figure 1.**
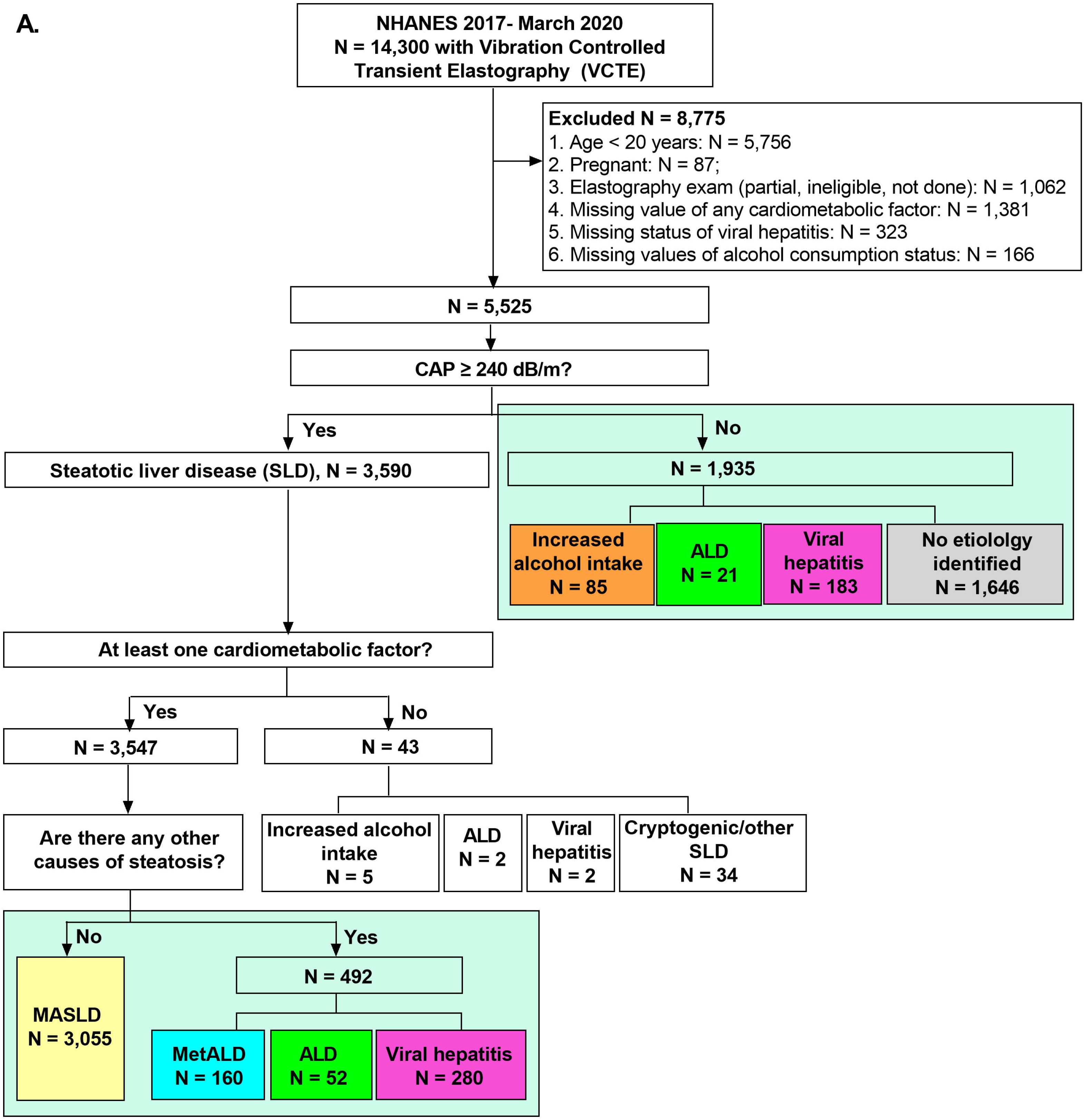
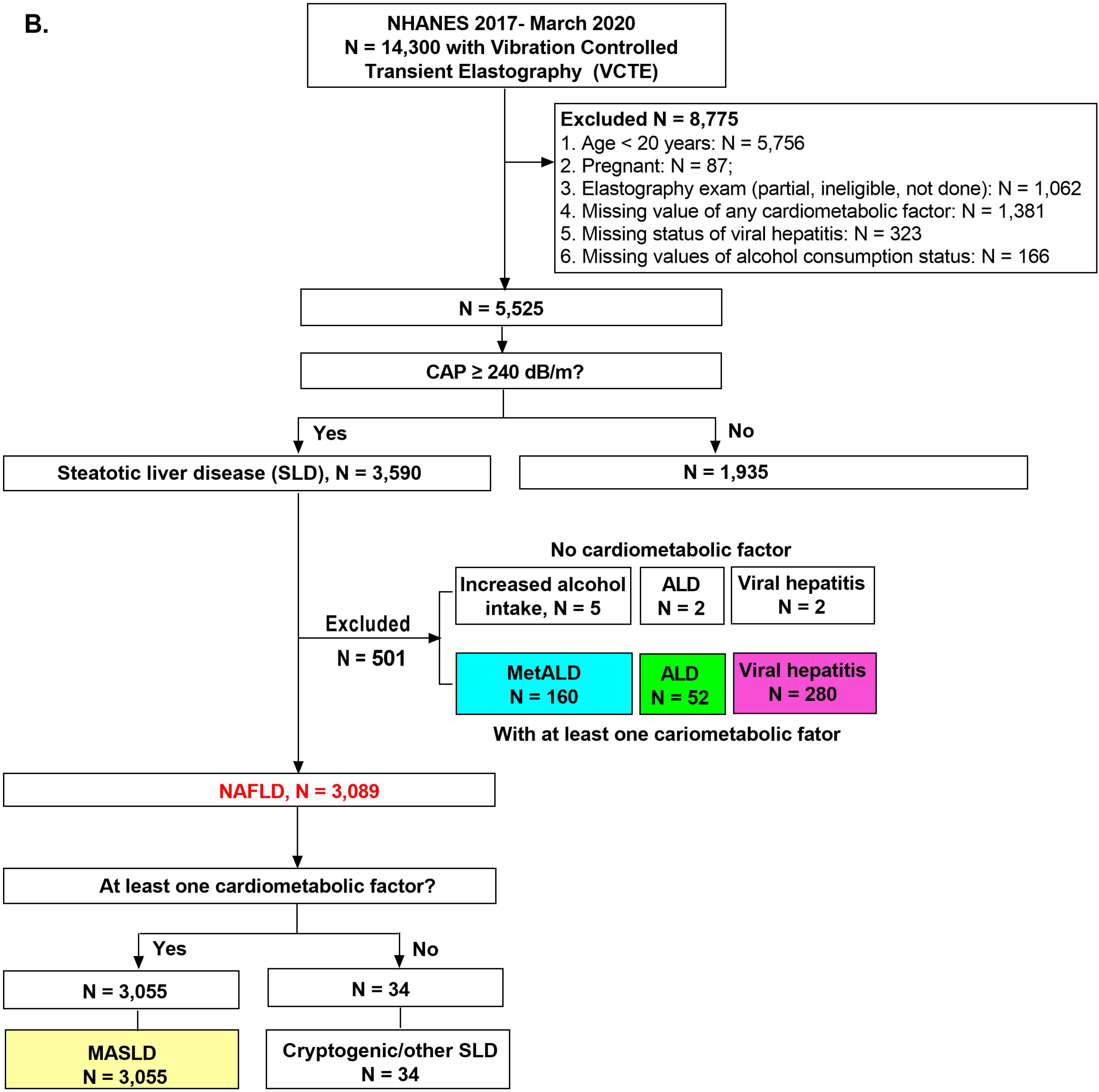
The new nomenclature’s diagnostic scheme applied to adults in the United States. The flowcharts show 5525 adults (age ≥ 20 years old) categorized **(A)** according to the proposed new nomenclature; **(B)** according to NAFLD criteria. Abbreviations: Alcohol associated liver disease, ALD; controlled attenuation parameter, CAP; National Health and Nutrition Evaluation Survey, NHANES; Metabolic dysfunction-associated alcohol-associated liver disease, MetALD; Metabolic dysfunction-associated liver disease, MASLD; Non-alcoholic fatty liver disease, NAFLD.

**Figure 2.**
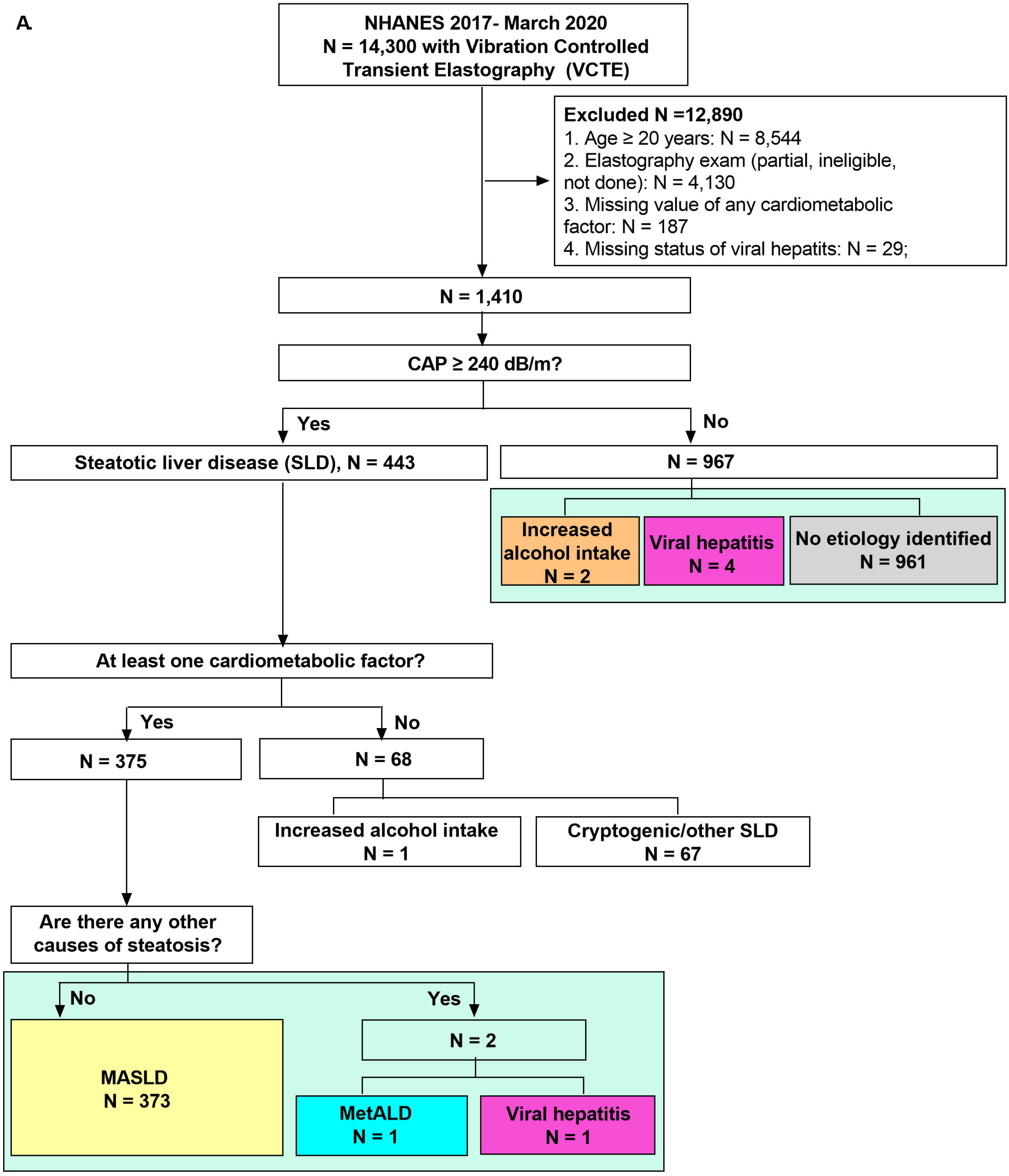
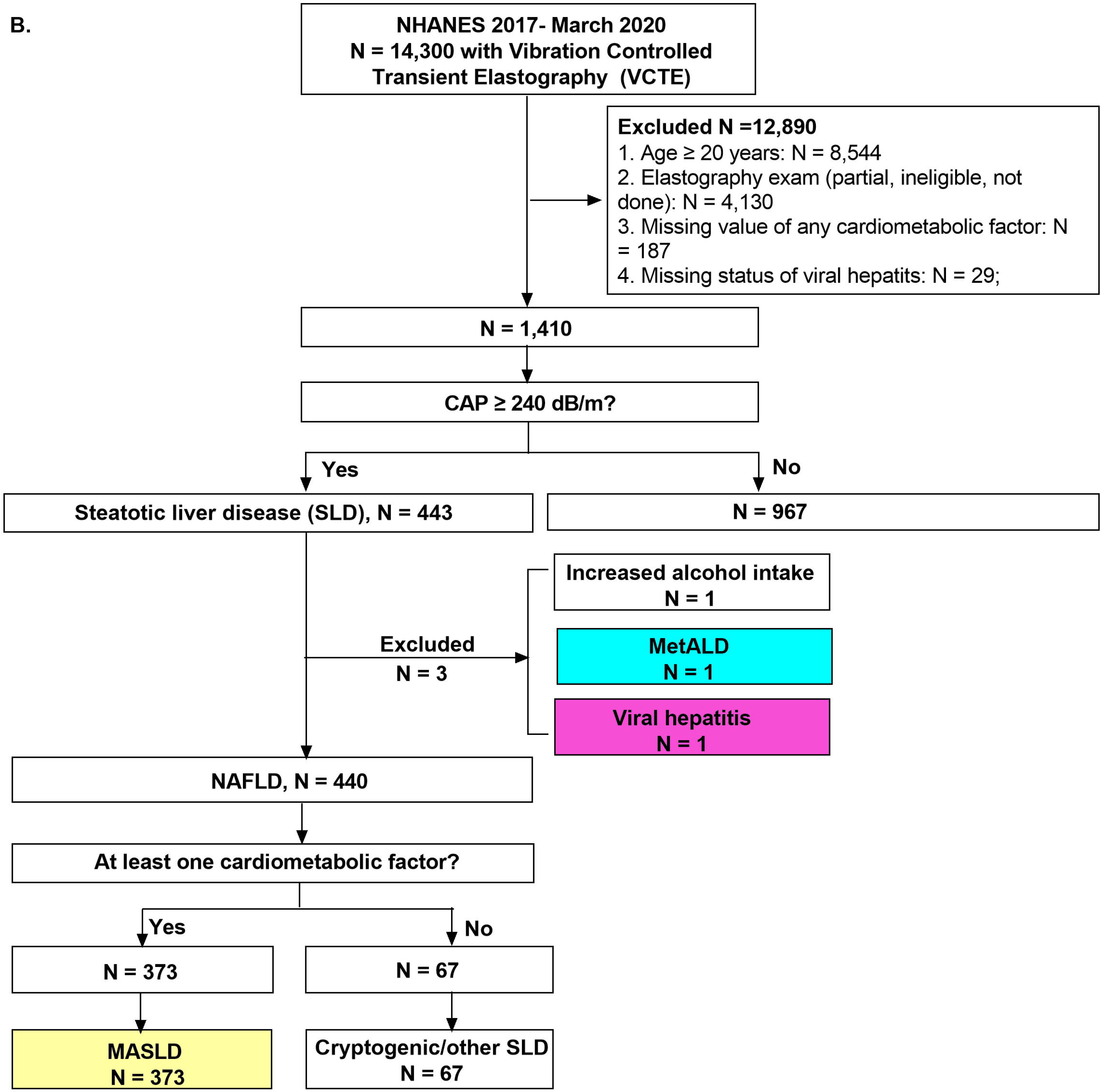
The new nomenclature’s diagnostic scheme applied to adolescents in the United States. The flowcharts show 1410 adolescents (age 12-19 years old) categorized **(A)** according to new nomenclature; **(B)** according to NAFLD criteria. Abbreviations: Alcohol associated liver disease, ALD; controlled attenuation parameter, CAP; National Health and Nutrition Evaluation Survey, NHANES; Metabolic dysfunction-associated alcohol-associated liver disease, MetALD; Metabolic dysfunction-associated liver disease, MASLD; Non-alcoholic fatty liver disease, NAFLD.

Figure 3 presents the age-standardized weighted prevalence of SLD with stratification by race/ethnicity during the years 2017-2020. Applying a VCTE-CAP threshold of 240 dB/m for the definition of steatosis grade 1 or higher, as recommended by the major supplier (EchoSens)^4^, the prevalence of SLD was 62.1% (95% CI, 59.8-64.3%) among adults and 30.5% (95% CI, 27.1-34.0%) among adolescents. Because opinions about the most appropriate VCTE-CAP threshold for defining SLD may vary, three different cut points were analyzed (Figure 3). At a threshold of 270 dB/m, 45.7% (95% CI, 44.0-47.4%) of adults and 16.5% (95% CI, 13.5-9.4%) of adolescents had SLD. Among adults, the prevalence of SLD was significantly lower in NHB than in NHW and it was higher in MA, regardless of the threshold. Among adolescents, the prevalence of SLD was similar in NHB and NHW, but higher in MA. Among MA adolescents, 45.6% (95% CI, 36.8-54.4%) had VCTE-CAP ≥ 240 dB/m and 30.6% (95% CI, 21.5, 39.7%) had VCTE-CAP ≥ 270 dB/m.

**Figure 3.**
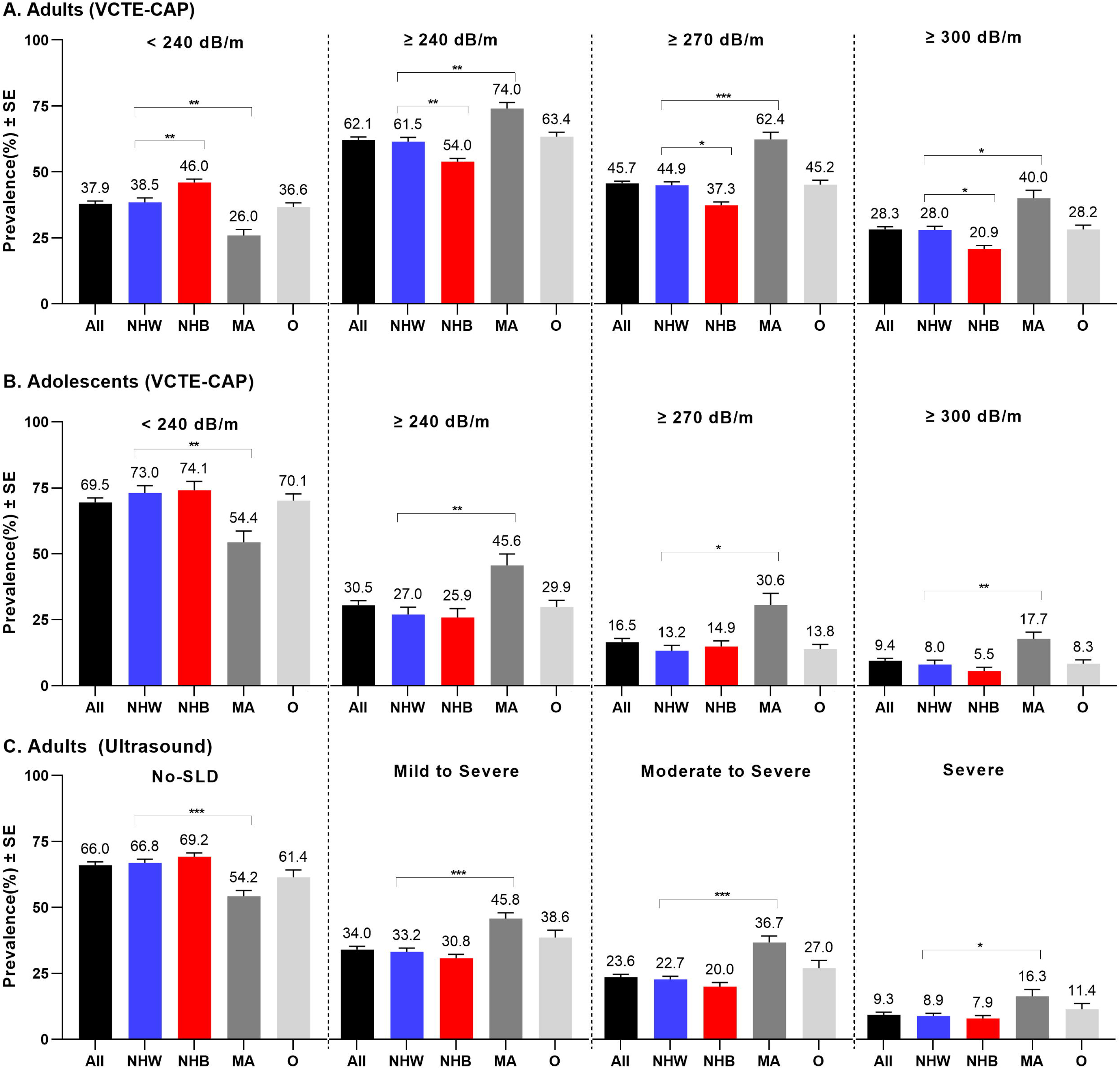
The age-standardized weighted percentage of SLD using different VCTE-CAP thresholds and abdominal ultrasound to define SLD, with stratification by race/ethnicity. The age-standardized weighted prevalence of SLD based on different cut points of VCTE-CAP thresholds and steatosis stages from abdominal ultrasound were determined for the total cohort (black), non-Hispanic White (NHW, blue), non-Hispanic Black (NHB, red), Mexican American (MA, grey) and other (O, light grey) racial/ethnic groups.**(A)** adults and **(B)** adolescents with VCTE data were divided by different VCTE-CAP thresholds as following: < 240 dB/m, ≥ 240 dB/m, ≥ 270 dB/m, and ≥ 300 dB/m. **(C)** adults with abdominal ultrasound data classified as No-SLD, mild to severe, moderate to severe and severe steatosis. Differences between groups were tested by univariate t statistic. Statistical significance was a two-sided P value <0.05. *P < 0.05, **P < 0.001, ***P < 0.0001. Abbreviations: Controlled attenuation parameter, CAP; steatotic liver diseases, SLD; vibration controlled transient elastography, VCTE; standard error, SE.

The second step in the diagnostic scheme divides people with SLD into those with and without at least one CMRF. Virtually all adults with SLD, 98.1% (95% CI, 97.1-99.1%), had at least one CMRF and 94.4% (95% CI, 92.8-96.0%) had increased adiposity, as determined by the composite endpoint of BMI ≥ 25 Kg/m^2^ and/or WC > 80 cm for women and 94 cm for men (Table 1). The vast majority of adults *<u>without</u>*SLD, 81.6% (95% CI, 78.3-84.9%), also had at least one CMRF, highlighting the large percentage of people without liver steatosis who have a CMRF. The prevalence of individual CMRFs varied by race/ethnicity in adults with and without SLD (Figure S1 and Figure S2). NHB persons had a higher prevalence of prediabetes/diabetes and a higher prevalence of elevated blood pressure than NHW in both SLD and No-SLD groups.

**Table 1.** Comparison of the age-standardized weighted prevalence of cardiometabolic risk factors in adults and adolescents with and without steatotic liver disease.

| Subjects | CMRF | Age-standardized weighted prevalence, % (95% CI) |  | Fold change* | P value |
| --- | --- | --- | --- | --- | --- |
|  |  | SLD | No-SLD |  |  |
| Adults | At least one CMRF | 98.1 (97.1, 99.1) | 81.6 (78.3, 84.9) | 1.2 | <0.0001 |
|  | Elevated adiposity | 94.4 (92.8, 96.0) | 64.7 (59.9, 69.5) | 1.5 | <0.0001 |
|  | Prediabetes/diabetes | 59.6 (56.7, 62.6) | 35.7 (32.0, 39.3) | 1.7 | <0.0001 |
|  | Elevated blood pressure | 45.3 (41.6, 49.1) | 30.7 (27.9, 33.5) | 1.5 | <0.0001 |
|  | Elevated triglycerides | 38.5 (36.1, 41) | 20.1 (17.4, 22.7) | 1.9 | <0.0001 |
|  | Reduced HDL | 53.9 (50.6, 57.1) | 31.6 (28.4, 34.8) | 1.7 | <0.0001 |
| Adolescents | At least one CMRF | 82.5 (77.3, 87.7) | 45.6 (40.5, 50.7) | 1.8 | <0.0001 |
|  | Elevated adiposity | 77.5 (72.4, 82.6) | 26.6 (22.2, 31.0) | 2.9 | <0.0001 |
|  | Prediabetes/diabetes | 27.7 (21.7, 33.7) | 18.6 (14.8, 22.4) | 1.5 | 0.003 |
|  | Elevated blood pressure | 2.1 (0.5, 3.8) | 1.6 (0.2, 3.1) | 1.3 | 0.6 |
|  | Elevated triglycerides | 5.1 (3.4, 6.7) | 0.9 (0.2, 1.7) | 5.7 | <0.0001 |
|  | Reduced HDL | 23.6 (19.1, 28) | 10.6 (7.7, 13.6) | 2.2 | <0.0001 |

Note: The difference of age-standardized weighted prevalence of CMRFs between SLD and No-SLD compared by univariate t statistic with significant level at $P < 0.05$ . \*Fold change was calculated by the prevalence in SLD/Non-SLD. Abbreviation: Cardiometabolic risk factor, CMRF; High-density lipoprotein cholesterol, HDL; Steatotic liver disease, SLD.

Nearly all adolescents with SLD, 82.5% (95%, 77.3-87.7%), had least one CMRF and 77.5% (95% CI, 72.4-82.6%) had elevated adiposity (Table 1). Almost half of adolescents without liver steatosis, 45.6% (95% CI, 40.5-50.7%), also had at least one CMRF (Table 1).

A major goal of the nomenclature review panel was to introduce a diagnostic scheme that would define a population with MASLD that recapitulated the population with NAFLD, keeping in mind that MASLD requires at least one CMRF and NAFLD does not. The goal of achieving equivalence was substantially met for adults with data from 2017-2020. Only about one percent of adults with NAFLD did not meet criteria for MASLD (Figure 1A and 1B), and all adults with NAFLD could be diagnosed with either definite MASLD or possible MASLD. About 15% of adolescents with NAFLD did not have a CMRF and thus did not meet criteria for MASLD (Figure 2A and 2B), but all could be diagnosed with MASLD or possible MASLD if all other causes of SLD could be ruled out. The diagnostic category “possible MASLD” spares patients and providers the confusion and anxiety that might result from being given the vague alternative diagnosis of cryptogenic SLD.

### Sensitivity analysis of the diagnostic scheme results

Sensitivity analysis was carried out on adults with abdominal ultrasound collected in NHANES III during the years 1988 to 1994.In this dataset, the age-standardized weighted prevalence of SLD was about 34% (Figure 3C), far lower than the 62.1% in the VCTE-CAP dataset collected during 2017-2020. Additionally, in the earlier dataset only about 92% of the population with NAFLD met the diagnostic criteria of MASLD (Figure S3A and Figure S3B). The lower SLD prevalence and the reduced concordance of NAFLD and MASLD likely reflect technical differences between VCTE-CAP^12^ and ultrasound, and changes in the population that occurred between the two data collection periods. As shown in Table S1, the prevalence of the composite endpoint of BMI ≥ 25 Kg/m^2^ and/or WC > 80 cm for women and 94 cm for men increased by over 15% between the two periods. These results suggest that estimates of the prevalence of SLD are likely to vary substantially depending on how intrahepatic lipid content is measured and scored, and on the prevalence of CMRFs in the population.

### Impact of the new nomenclature on referrals and on the hepatology workforce

The American Gastroenterological Association (AGA) recommends that adults in the community with VCTE-defined liver stiffness ≥ 8.0 kPa be referred to hepatology^13^. These criteria were applied to the NHANES dataset. The estimated average adult and adolescent populations of the United States at the time of the NHANES survey in 2017-2020 were 240.4 million and 33.8 million^10^, respectively.

At a threshold VCTE-CAP of 240 dB/m, 153.3 million (95% CI, 147.7-158.9) adults had SLD. Among the SLD group, a total of 19.3 million (95% CI, 15.8-22.8) had liver stiffness ≥ 8.0 kPa and would qualify for referral to hepatology, including 16.3 million (95% CI, 13.2-19.3) with MASLD. When a higher threshold (VCTE-CAP ≥ 270 dB/m) was used to define SLD, a total of 17.8 million (95% CI, 14.2-21.3) adults with SLD met criteria for hepatology referral, including 14.9 million (95% CI, 11.9-18.0) with MASLD. Regardless of the threshold used to define liver steatosis, a very large number of adults meet criteria for referral to hepatology, creating an increased demand for liver care services.

A total of 16.1 million (95% CI, 12.9-19.2) adults with SLD had VCTE ≥ 8.6 kPa, the threshold for significant (≥ F2) fibrosis^5^, including 13.5 million (95% CI, 10.9-16.2) with MASLD. Using the 270 dB/m threshold to define steatosis, a total of 15.0 million (95% CI, 11.7, 18.3) adults with SLD had a high risk of significant fibrosis (Table 2).

**Table 2.** Estimated population counts of U.S adults with steatotic liver disease meeting criteria for referral to hepatology and the counts with significant fibrosis.

| SLD category |  | Weighted %<br>(95% CI) in total<br>population | Population<br>estimation of each<br>SLD category<br>(million) | Weighted % (95%<br>CI) of LS ≥ 8.0<br>kPa in each SLD<br>category | Population<br>estimation of LS ≥<br>8.0 kPa in each LD<br>category (million) | Weighted %<br>(95% CI) of LS ≥<br>8.6 kPa in each<br>SLD category | Population<br>estimation of LS<br>≥ 8.6 kPa in each<br>LD category<br>(million) |
| --- | --- | --- | --- | --- | --- | --- | --- |
| **SLD (CAP ≥ 240 dB/m)** |  | 63.7 (61.4, 66.1) | 153.3 (147.7, 158.9) | 12.6 (10.3, 14.8) | 19.3 (15.8, 22.8) | 10.5 (8.4, 12.5) | 16.1 (12.9, 19.2) |
| **Sub-**<br>**categories**<br>**of SLD**<br>**(CAP ≥**<br>**240 dB/m)** | **MASLD** | 54.7 (51.8, 57.7) | 131.6 (124.6, 138.7) | 12.4 (10.0, 14.7) | 16.3 (13.2, 19.3) | 10.3 (8.2, 12.3) | 13.5 (10.9, 16.2) |
|  | **MetALD** | 4.0 (3.2, 4.7) | 9.5 (7.7, 11.4) | 10.5 (2.9, 18.1) | 1.0 (0.3, 1.7) | 9.9 (2.4, 17.4) | 0.9 (0.2, 1.7) |
|  | **ALD + CMRF** | 0.9 (0.4, 1.4) | 2.2 (1, 3.3) | 20.0 (6.6, 33.4) | 0.4 (0.1, 0.7) | 14.4 (1.0, 27.7) | 0.3 (0.02, 0.6) |
|  | **VH + CMRF** | 3.2 (2.2, 4.3) | 7.8 (5.3, 10.3) | 19.1 (11.9, 26.4) | 1.5 (0.9, 2.1) | 15.8 (8.8, 22.7) | 1.2 (0.7, 1.8) |
|  | **No CMRF** | 0.9 (0.4, 1.4) | 2.2 (1.1, 3.3) | 2.5 (0, 7.8) | 0.06 (0, 0.2) | 2.5 (0, 7.8) | 0.06 (0, 0.2) |
| **SLD (CAP ≥ 270 dB/m)** |  | 47.1 (45.3, 49.0) | 113.4 (109.0, 117.7) | 15.7 (12.6, 18.8) | 17.8 (14.2, 21.3) | 13.3 (10.3, 16.2) | 15.0 (11.7, 18.3) |
| **Sub-**<br>**categories**<br>**of SLD**<br>**(CAP ≥**<br>**270 dB/m)** | **MASLD** | 40.8 (38.5, 43.1) | 98.1 (92.7, 103.5) | 15.2 (12.1, 18.3) | 14.9 (11.9, 18.0) | 12.9 (10.0, 15.7) | 12.6 (9.8, 15.4) |
|  | **MetALD** | 2.9 (2.3, 3.5) | 7.0 (5.5, 8.5) | 14.4 (4.5, 24.2) | 1.0 (0.3, 1.7) | 13.5 (3.7, 23.3) | 0.9 (0.3, 1.6) |
|  | **ALD + CMRF** | 0.7 (0.3-1.2) | 1.7 (0.7, 2.8) | 23.9 (9.9, 37.9) | 0.4 (0.2, 0.6) | 16.9 (2.0, 31.8) | 0.3 (0.03, 0.5) |
|  | **VH + CMRF** | 2.5 (1.6, 3.4) | 6.0 (3.8, 8.2) | 22.6 (13.0, 32.3) | 1.4 (0.8, 1.9) | 18.7 (9.9, 27.5) | 1.1 (0.6, 1.7) |
|  | **No CMRF** | 0.2 (0.04, 0.39) | 0.5 (0.09, 0.9) | 10.7 (0, 32.2) | 0.05 (0, 0.2) | 10.7 (0, 32.2) | 0.05 (0, 0.2) |

Abbreviation: alcoholic associated liver disease, ALD; cardiometabolic risk factor, CMRF; controlled attenuation parameter, CAP; Viral hepatitis, VH; metabolic dysfunction associated liver disease, MASLD; Liver stiffness, LS; Steatotic liver disease, SLD;

The age-adjusted weighted percentage of adults with SLD who reported being aware of having a liver disease was only 5.3% (95% CI, 1.3-6.3%). Among adults with SLD and liver stiffness ≥ 8.0 kPa, only 12.2% (95% CI, 7.0-17.3%) reported being aware and among adults with SLD and liver stiffness ≥ 8.6 kPa, only 13.5% (95% CI, 7.4-19.6%) reported being aware (Table S2).

These results show that although self-reported awareness of having a liver disease was higher among adults with fibrosis than among those without (Figure S4), awareness was generally very low. These findings highlight the need for increased patient awareness of SLD, which is a problem the new nomenclature and diagnostic scheme are designed to address.

At a threshold VCTE-CAP of 240 dB/m, 10.3 million (95% CI, 9.1-11.5) adolescents had SLD, including 8.5 million (95% CI, 7.4-9.6) with MASLD and 1.7 million (95% CI, 1.2-2.3) with No-CMRF/Cryptogenic/other SLD. Among adolescents with SLD, 1.0 million (95% CI, 0.3-1.8) had fibrosis liver stiffness ≥ 7.4 kPa^7^ and 0.7 million (95% CI, 0.2-1.2) had liver stiffness ≥ 8.0 kPa (Table S3). When VCTE-CAP ≥ 270 dB/m was used to define SLD, 5.5 million (95% CI, 4.6-6.5) adolescents had SLD, including 5.1 million (95% CI, 4.2-6.0) with MASLD and 0.8 million (95% CI, 0.3-1.3) with increased liver stiffness (Table S3). These numbers show that a very large number of adolescents are likely in need of referral to hepatology.

### Risk factors for significant fibrosis

Because fibrosis is an important determinant of liver-related outcomes, including mortality^14, 15^, the liver diseases defined by the new and more precise diagnostic scheme were ranked based on their population-level contribution to significant fibrosis, defined as VCTE liver stiffness ≥ 8.6 kPa, the threshold for stage F2 fibrosis and higher^5^. VCTE-CAP ≥ 240 dB/m was used to define steatosis. The SLD group accounted for 87.6% (95% CI, 84.6-90.6%) of the population at high risk for ≥ F2 fibrosis, while the No-SLD group accounted for 12.4% (95% CI, 9.4-15.4%) (Figure 4A). MASLD accounted for the largest percentage, 73.7% (95% CI, 67.8-79.7%). Surprisingly, the No Etiology Identified group (NEI), which lacked SLD or any other identified cause of LD in the NHANES dataset, accounted for the second highest percentage, 8.7% (95% CI, 5.6-11.7%), and viral hepatitis/SLD/CMRF was the third highest, 6.7% (95% CI, 3.9-9.5%) (Figure 4B).

**Figure 4.**
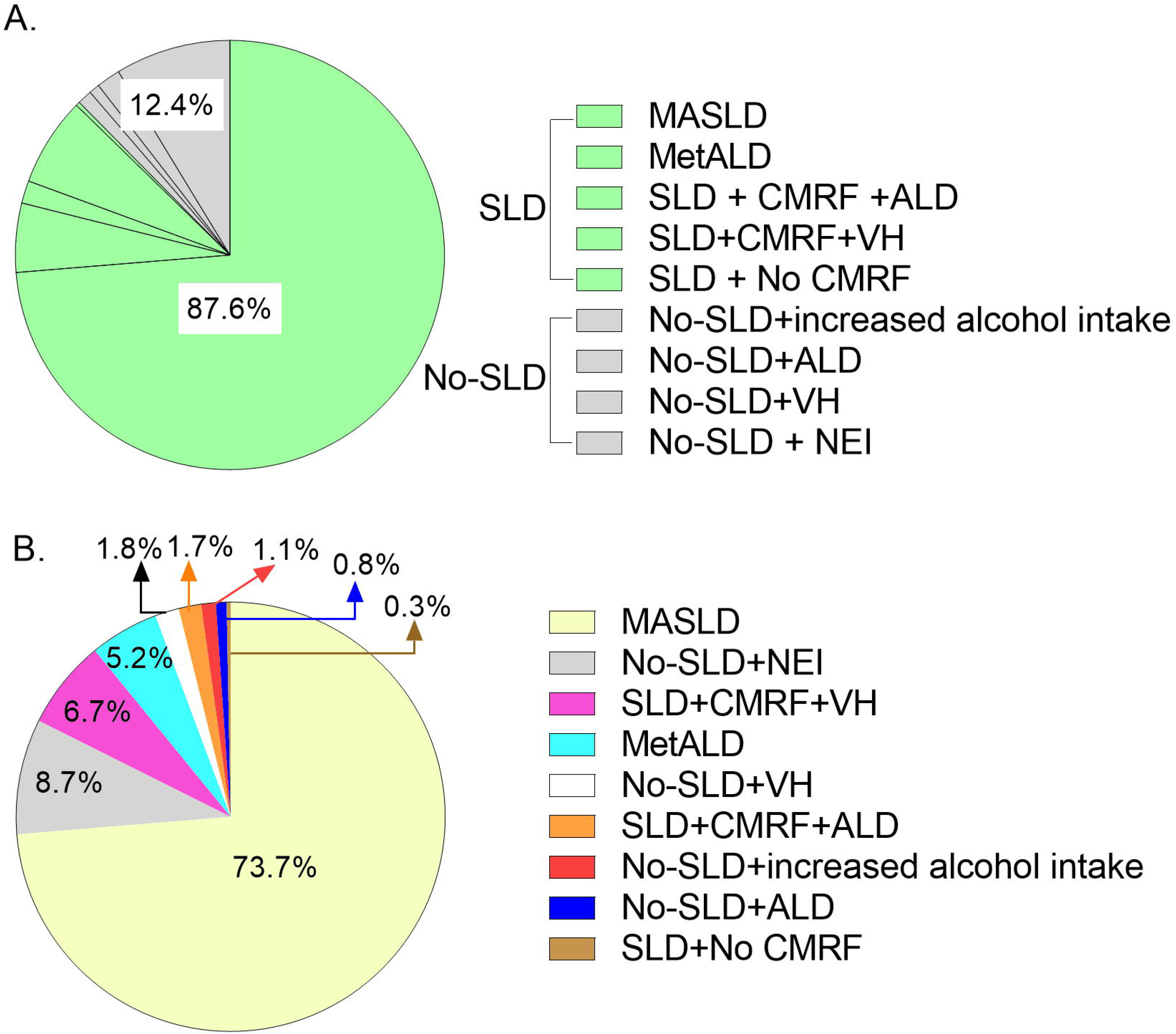
The population-level contribution of various liver diseases to significant fibrosis (≥ F2) in adults. Significant fibrosis was defined by VCTE liver stiffness ≥ 8.6 kPa. The distribution of significant fibrosis is shown in pie charts with the weighted percentage of significant fibrosis. **(A)** grouped as SLD and non-SLD; **(B)** liver diseases sorted by weighted percentage values of significant fibrosis. Abbreviations: Alcohol associated liver disease, ALD; cardiometabolic risk factors, CMRF; metabolic dysfunction-associated alcohol-associated liver disease, MetALD; Metabolic dysfunction-associated liver disease, MASLD; steatotic liver disease; SLD; viral hepatitis, VH; No etiology identified, NEI.

Risk factors associated with liver fibrosis ≥ F2 were investigated by logistic regression. Variables with P values < 0.1 in univariable models (Table S4) were included in multivariable models, as described in Methods. Among patients with MASLD, fibrosis risk factors varied strikingly by race (Table 3). Among NHB, diabetes was not associated with ≥ F2 fibrosis, odds ratio (OR) =1.07 (95% CI, 0.54-2.13), whereas among NHW, MA, and O, diabetes increased the odds dramatically, with ORs of 3.05 (95% CI, 1.82-5.11), 3.47 (95% CI, 1.29, 9.33), and 2.51 (95% CI, 1.33-4.72), respectively. Similarly, hypertension was not associated with ≥ F2 fibrosis in NHB, but had an OR of 1.92 (95% CI, 1.02-3.62) in NHW. Conversely, among NHW, past and current alcohol consumption greatly reduced the odds of ≥ F2 fibrosis compared to lifetime abstainers whereas this association was not observed for NHB, MA, and O.

**Table 3.** Odds ratios of factors associated with significant fibrosis (LS ≥ 8.6 kPa) among adults with MASLD in multivariable logistic regression.

|  | OR (95%CI) |  |  |  |  |
| --- | --- | --- | --- | --- | --- |
|  | Total | NHW | NHB | MA | Other |
| **Sample size** | **3,055** | **1,180** | **674** | **428** | **773** |
| **Significant fibrosis** | **331** | **138** | **73** | **49** | **71** |
| **Age (unit 10 years)** | 1.00 (0.89, 1.13) | 0.93 (0.79, 1.10) | **1.46 (1.17, 1.81)\*\*** | **1.34 (1.03, 1.75)\*** | 1.03 (0.79, 1.34) |
| **Diabetes** |  |  |  |  |  |
| No | Reference |  |  |  |  |
| Yes | **2.77 (1.94, 3.96)‡** | **3.05 (1.82, 5.11)‡** | 1.07 (0.54, 2.13) | **3.47 (1.29, 9.33)\*** | **2.51 (1.33, 4.72)\*** |
| **Hypertension** |  |  |  |  |  |
| No | Reference |  |  |  |  |
| Yes | **1.75 (1.22, 2.51)\*** | **1.92 (1.02, 3.62)\*** | 1.32 (0.40, 4.35) | 1.12 (0.32, 3.94) | 1.77 (0.78, 4.00) |
| **Waist circumference (cm)** | **1.07 (1.06, 1.08)‡** | **1.07 (1.05, 1.08)‡** | **1.07 (1.04, 1.10)‡** | **1.07 (1.03, 1.12)†** | **1.08 (1.06, 1.10)‡** |
| **Alcohol use** |  |  |  |  |  |
| Lifetime abstainers | Reference |  |  |  |  |
| Former drinkers | **0.45 (0.23, 0.88)\*** | **0.30 (0.11, 0.80)\*** | 1.18 (0.27, 5.17) | 0.63 (0.20, 1.99) | 1.05 (0.29, 3.80) |
| Current drinkers | **0.48 (0.29, 0.80)\*** | **0.32 (0.15, 0.67)\*** | 1.23 (0.34, 4.44) | 1.20 (0.33, 4.41) | 0.77 (0.30, 1.98) |

Note: multiple imputation was performed in univariable and multivariable logistic regression models. Variables with P value < 0.1 in the univariable analysis in the in total cohort were included in multivariable models. \*P < .05, †P < .001, ‡P < .0001.
Abbreviations: odds ratio, OR; confidence interval, CI; Non-Hispanic White, NHW; Non-Hispanic Black, NHB; Mexican American, MA; metabolic dysfunction-associated liver disease, MASLD; other race, O.

The proposed precision nomenclature defined a new subgroup, MetALD, in which both metabolic dysfunction and alcohol exposure may be driving liver disease progression. Interestingly, ≥ F2 fibrosis risk did not differ significantly between the MASLD and MetALD subgroups (Figure S5) but was higher in the viral hepatitis/SLD/CMRF subgroup than in the MASLD subgroup, OR = 1.64 (95% CI, 1.01-2.66) with adjustment for age, sex and race/ethnicity (Figure S5 and Figure S6). Within viral hepatitis subgroups, ≥ F2 fibrosis risk was higher in the group with SLD, OR = 2.68 (95% CI, 1.15-6.21) with adjustment for age (Figure S7).

In the No Etiology Identified subgroup (which lacked SLD or any other identified cause of LD), high (4^th^ quartile) blood levels of lead (Pb), OR = 4.84 (95% CI, 2.48-9.43), and cadmium (Cd), OR = 1.91 (95% CI, 1.10-3.32), were associated with ≥ F2 fibrosis in univariable analysis, but diabetes was not, OR = 1.32 (95% CI, 0.52-3.32; p = 0.6) (Table S5). In the multivariable model, Pb remained significantly associated with ≥ F2 fibrosis, with an OR of 3.89 (95% CI, 2.00-7.56). Male sex and elevated triglycerides were also significant, but the ORs were much lower than Pb (Table 4).

**Table 4.** Odds ratios of factors associated with significant fibrosis (LS≥ 8.6 kPa) among adults with No Etiology Identified (no SLD or any other identified LD)

|  | OR (95%CI) |
| --- | --- |
| **Sample size** | **1,646** |
| **Significant fibrosis** | **40** |
| **Gender** |  |
| Female | Reference |
| Male | **2.12 (1.10, 4.10)\*** |
| **Waist circumference (cm)** | 1.04 (0.99, 1.09) |
| **Elevated triglycerides** |  |
| No | Reference |
| Yes | **1.63 (1.01, 2.62)\*** |
| **Lead** |  |
| Q1-3 | Reference |
| Q4 | **3.89 (2.00, 7.56)†** |

Multiple imputation was performed in univariable and multivariable logistic regression models. Variables with P value < 0.1 in the univariable analysis in the total cohort were included in multivariable models. \*P < .05, †P < .001. Abbreviations: odds ratio, OR; confidence interval, CI; quartile, Q.

### Mortality risk

The NHANES III (1988-1994) dataset provides the information needed to evaluate mortality risk among people in the liver disease subgroups defined by the new diagnostic criteria (Figure S3). Data were linked to the National Death Index to assess mortality through December 31, 2019, providing about 30 years of follow-up. Compared to the No-SLD group, the SLD group did not have significantly increased all-cause mortality, hazard ratio (HR) = 1.07 (95% CI, 0.98-1.18), heart disease-related mortality, or cancer-related mortality after adjusting for covariates (Table S6). Additionally, mortality rates were similar in the No-SLD group and the SLD/CMRF group (Table S7). Within the SLD/CMRF group, the viral hepatitis and ALD subgroups had significantly higher risk of all-cause mortality than the MASLD group, with HR = 1.39 (95% CI, 1.01-1.91) and HR = 1.99 (95% CI, 1.29-3.06), respectively (Table S8). There were no significant differences in all-cause mortality in a series of additional analyses that compared a) the MetALD and No-SLD/Increased alcohol subgroups (Table S9); b) the viral hepatitis/No-SLD and the viral hepatitis/SLD/CMRF subgroups (Table S10); c) the ALD/No-SLD and ALD/SLD/CMRF subgroups (Table S11); and d) the No Etiology Identified/No-SLD and MASLD subgroups (Table S12).

## Discussion

This study strengthens the rationale for the “call for unity” in steatotic liver disease nomenclature^1, 2^. It projects the new nomenclature’s diagnostic algorithm onto the housed civilian population of the United States^1, 3^ The findings clarify how successfully the new nomenclature achieved its goals of a) preserving the NAFLD patient group under a new name, MASLD, and b) helping patients and providers understand the main drivers of liver injury.

A major contribution of the new nomenclature is that it provides a framework for investigating the importance of liver steatosis as a driver of disease progression in patients with all types of liver disease, e.g., viral hepatitis. The first branch point in the diagnostic algorithm is the division between patients with and without liver steatosis. This study showed that the viral hepatitis/SLD subgroup had a higher risk of ≥ F2 fibrosis than the viral hepatitis/No-SLD subgroup, illustrating the value of having an overarching term, SLD, which provides a formal structure for evaluating the clinical ramifications of hepatic steatosis and its drivers. By making steatosis a diagnostic consideration across all liver diseases, the new nomenclature and diagnostic algorithm will likely increase demand for the non-invasive assessment of intrahepatic lipids. Meeting this demand may require additional resources and will benefit from clear guidelines about the diagnostic methods and threshold values that should be used to define steatosis. Additionally, given the importance the new diagnostic algorithm places on steatosis, it will be important to ensure that SLD does lead to a diagnosis MASLD until a full hepatology work up has been completed.

This investigation demonstrated that the new nomenclature achieved nearly complete overlap between NAFLD and MASLD for adults in the NHANES 2017-2020 cohorts, accomplishing one of its main goals. In this group, over 98% of the people with SLD and no other identifiable cause of liver disease--*i.e*., those with NAFLD--had at least one of the five CMRFs designated by the nomenclature panel and thus met the diagnostic criteria of MASLD. These results are consistent with those reported from an international cohort^1^. A high concordance between MASLD and NAFLD was virtually assured by the fact that over 90% of adults in the United States have at least one CMRF. The designated CMRFs are widely available in medical records and can be measured accurately, justifying their selection as diagnostic criteria, however because their prevalence is so high, they have limited power to stratify the population based on liver disease risk. The extremely high prevalence of CRMFs is concerning. The new nomenclature makes a valuable contribution by calling attention to this important personal and public health problem.

The study established that over 62% of adults and 30% of adolescents have SLD. Given these percentages, it is not surprising that an enormous number of adults with SLD, around 19 million, qualify for referral to hepatology according to AGA guidelines. This far exceeds the capacity of the hepatology workforce. If SLD awareness increases as a consequence of the nomenclature changes, as anticipated by the nomenclature panel, either referral guidelines will need to be changed or the workforce will need to expand very rapidly.

This study identified two populations in which the overlap of NAFLD and MASLD was less complete than in adults in the 2017-2020 cohorts: Adolescents in the 2017-2020 cohorts and adults with ultrasound defined SLD whose data were collected in 1988-1994, at a time when obesity and other CMRFs were less prevalent than in later years. These results raise the possibility that a patient’s diagnosis may vary depending on the sensitivity of the modality used to measure intrahepatic lipids (VCTE-CAP is more sensitive than ultrasound) and on the prevalence of CMRFs in the population. It will be interesting to see if improvements in obesity treatments increase the number of people in the non-concordant category who have SLD and no CMRF (and no other cause of liver disease) and to learn the natural history of their liver disease. Under the new nomenclature, these patients will receive the less-than-ideal diagnosis of cryptogenic SLD or possible MASLD. As understanding of MASLD pathogenesis deepens and biomarkers are approved, more definitive and stable diagnostic criteria will likely emerge. Similarly, research may change the diagnostic criteria of burnt-out NASH/MASH^16–18^, a term that may be applied to patients with cryptogenic cirrhosis according to the new nomenclature even if they lack the hallmark feature of SLD—liver steatosis.

A second major goal of the nomenclature panel was to help patients and providers understand the main drivers of their liver disease. To this end, they sought a diagnostic term that “describes the underlying cause of the disease” and wanted criteria that identify “individuals with obesity and cardiometabolic risk factors in the context or regional/ethnic differences”^1^. Ideally, the panel sought CMRFs that would be race-neutral and help patients understand the causal relationship between metabolic dysfunction, particularly insulin resistance, and disease progression. It is thus noteworthy that MASLD-fibrosis was not associated with diabetes or hypertension in NHB in this study, consistent with earlier findings ^8, 19, 20^. These results highlight the need for a better understanding of the relationship between metabolic parameters and liver disease progression, especially in NHB.

Interestingly, the impact of alcohol consumption on liver fibrosis risk in adults with MASLD also differed between racial and ethnic groups. Among NHW, lifetime abstainers had a higher risk of significant liver fibrosis than current and former consumers of alcohol, but this apparent protective effect was not observed in other groups. Among all adults (without stratification by race/ethnicity), there was no difference in fibrosis risk between the MASLD and the MetALD groups. These findings will add to the debate about the riskiness of moderate alcohol consumption^21, 22^. They highlights the need to employ objective assays for alcohol exposure, such as phosphatidylethanol testing^23^ and to redouble efforts to uncover possible confounders in the relationship between alcohol consumption and fibrosis. Longitudinal studies will be required to determine the implications of the new diagnostic criteria for alcohol-related liver disease progression.

Alcohol is only one of a very large number of hepatotoxic substances. The increasing evidence that environmental toxins are associated with SLD^24^ and data showing the hepatoxic effects of heavy metals prompted an analysis of Pb and Cd as possible fibrosis risk factors. Pb was strongly associated with liver fibrosis (OR = 3.89) in people without SLD and with no other identified cause of LD. The toxic metals were not associated with fibrosis risk in adults with MASLD.

This study has strengths and weaknesses. The strengths include the use of two non-overlapping NHANES datasets that provide nationally representative information, the use of VCTE, a widely-used methodology to estimate steatosis and fibrosis in the main analysis, and the analysis of mortality data with about 30 years of follow up. The limitations include the cross-sectional design of the NHANES study, the lack of biopsy, MRI or MR elastography data to confirm liver steatosis and liver fibrosis, and the resulting inability to evaluate metabolic dysfunction-associated steatohepatitis (MASH). Additionally, NHANES has limited data about the less common liver diseases, such as autoimmune hepatitis; individuals with these diseases may be misclassified.

## Conclusions

The precise and patient-centric nomenclature proposed by a recent consensus panel of experts highlights the extraordinary prevalence of SLD and provides a non-stigmatizing alternative to NAFLD. The diagnostic criteria for MASLD, the replacement term, capture nearly the same patient population and thereby preserve pharmaceutical and biomarker development pipelines. Risk factors of MASLD-fibrosis vary by race and do not include diabetes or hypertension in non-Hispanic Black individuals. This suggests that the pathogenic pathways connecting liver fibrosis to the markers of metabolic dysfunction selected by the nomenclature committee vary by race, which is problematic. Pb exposure contributed to the risk of fibrosis in adults without SLD or any other identified cause of liver disease, highlighting the importance of environmental liver toxins as drivers of cryptogenic liver disease. This nationally-representative study establishes that new and more precise nomenclature provides a framework for understanding liver disease pathogenesis and has utility for public health and workforce planning.

## Data Availability

All data produced are available online at https://www.ncdc.gov/nchs/nhanes/continuousnhanes/default.aspx?cycle=2017-2020

## Acknowledgements

The authors thank Dr. Scott Friedman for sharing his insights and careful reading of the manuscript.

## List of Abbreviations

Alcohol associated liver disease (ALD)

Body mass index (BMI)

Cardiometabolic risk factors (CMRF)

Confidence interval (CI)

Controlled attenuation parameter (CAP)

Diastolic blood pressure (DBP)

Fasting plasma glucose (FPG)

Hepatitis B virus (HBV)

Hepatitis C virus (HCV)

High-density lipoprotein cholesterol (HDL)

Metabolic dysfunction-associated alcohol-associated liver disease (MetALD)

Metabolic dysfunction-associated liver disease (MASLD)

Mexican American (MA)

National Health and Nutrition Examination Survey (NHANES)

No etiology identified (NEI)

Non-alcoholic fatty liver disease (NAFLD)

Non-Hispanic Black (NHB)

Non-Hispanic White (NHW)

Odds ratio (OR)

Steatotic liver diseases (SLD)

Systolic blood pressure (SBP)

Vibration controlled transient elastography (VCTE)

Viral hepatitis (VH)

Waist circumference (WC)

