## Supplementary material for "Much more than a name change: Impact of the new steatotic liver disease nomenclature on clinical algorithms and disease classification in U.S. adults and adolescents": Figure S, Table S

**Table content**

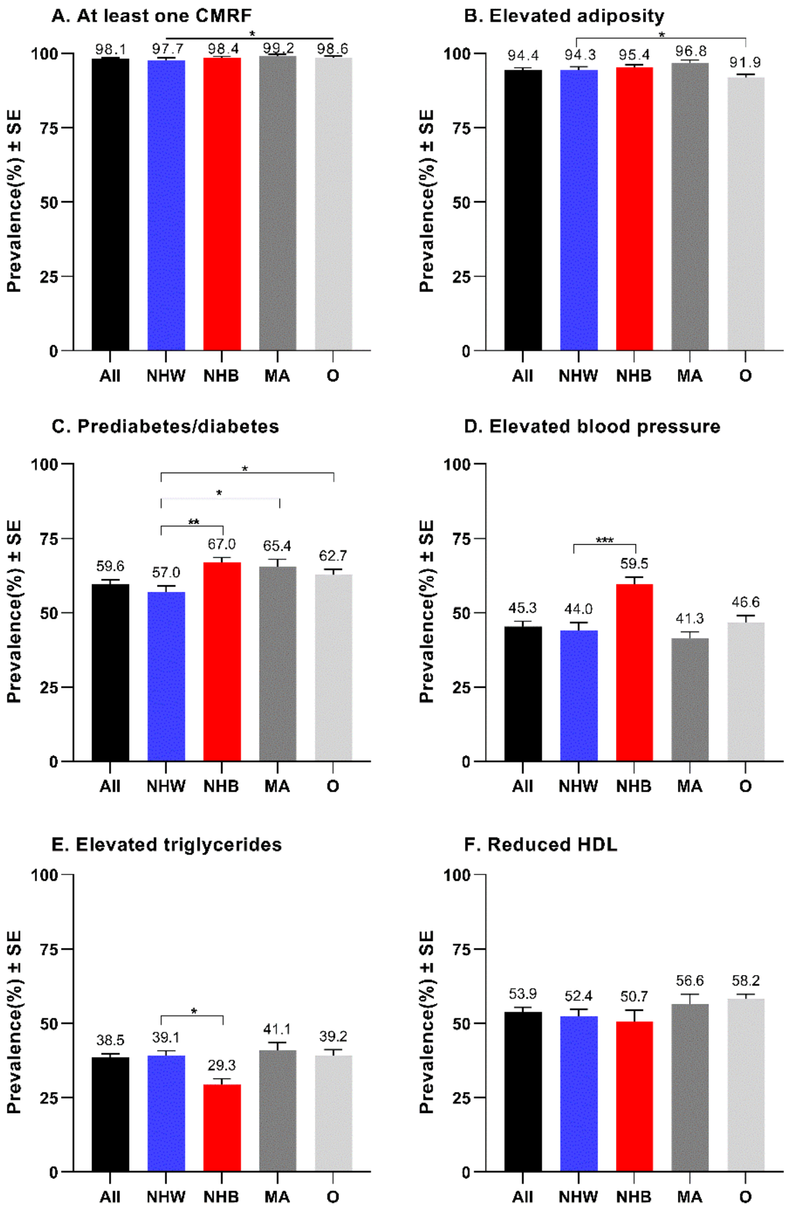

**Figure S1. Age-standardized weighted prevalence of cardiometabolic risk factors among adults with SLD stratified by race/ethnicity.** The age-standardized weighted prevalence of cardiometabolic risk factors (CMRFs) were determined in the total cohort (black), non-Hispanic White (NHW, blue), non-Hispanic Black (NHB, red), Mexican American (MA, grey) and other (O, light grey) racial/ethnic groups. (A) at least one CMRF; (B) elevated adiposity; (C) prediabetes/diabetes; (D) elevated blood pressure; (E) elevated triglycerides; (F) reduced HDL. Differences between groups were tested by univariate t statistic. *$P < 0.05$, **$P < 0.001$, ***$P < 0.0001$. Abbreviations: High-density lipoprotein cholesterol, HDL; steatotic liver disease, SLD; standard error, SE.

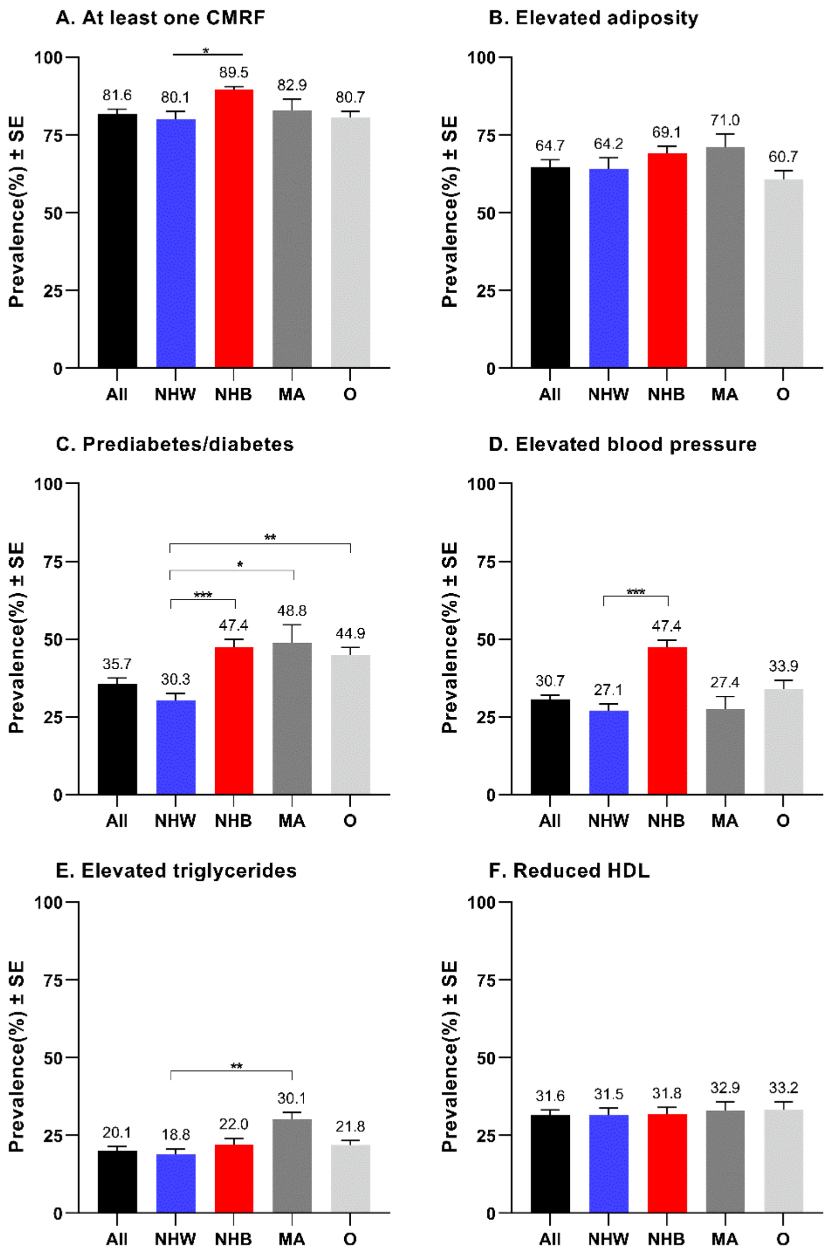

**Figure S2. Age-standardized weighted prevalence of cardiometabolic risk factors among adults in the No-SLD category, stratified by race/ethnicity.** The age-standardized weighted prevalence of cardiometabolic risk factors (CMRFs) were determined in the total cohort (black), non-Hispanic White (NHW, blue), non-Hispanic Black (NHB, red), Mexican American (MA, grey) and other (O, light grey) racial/ethnic groups. (A) at least one CMRF; (B) elevated adiposity; (C) prediabetes/diabetes; (D) elevated blood pressure; (E) elevated triglycerides; (F) reduced HDL. Differences between groups were tested by univariate t statistic. *P < 0.05, **P < 0.001, ***P < 0.0001. Abbreviations: High-density lipoprotein cholesterol, HDL; steatotic liver disease, SLD; standard error, SE.

A.

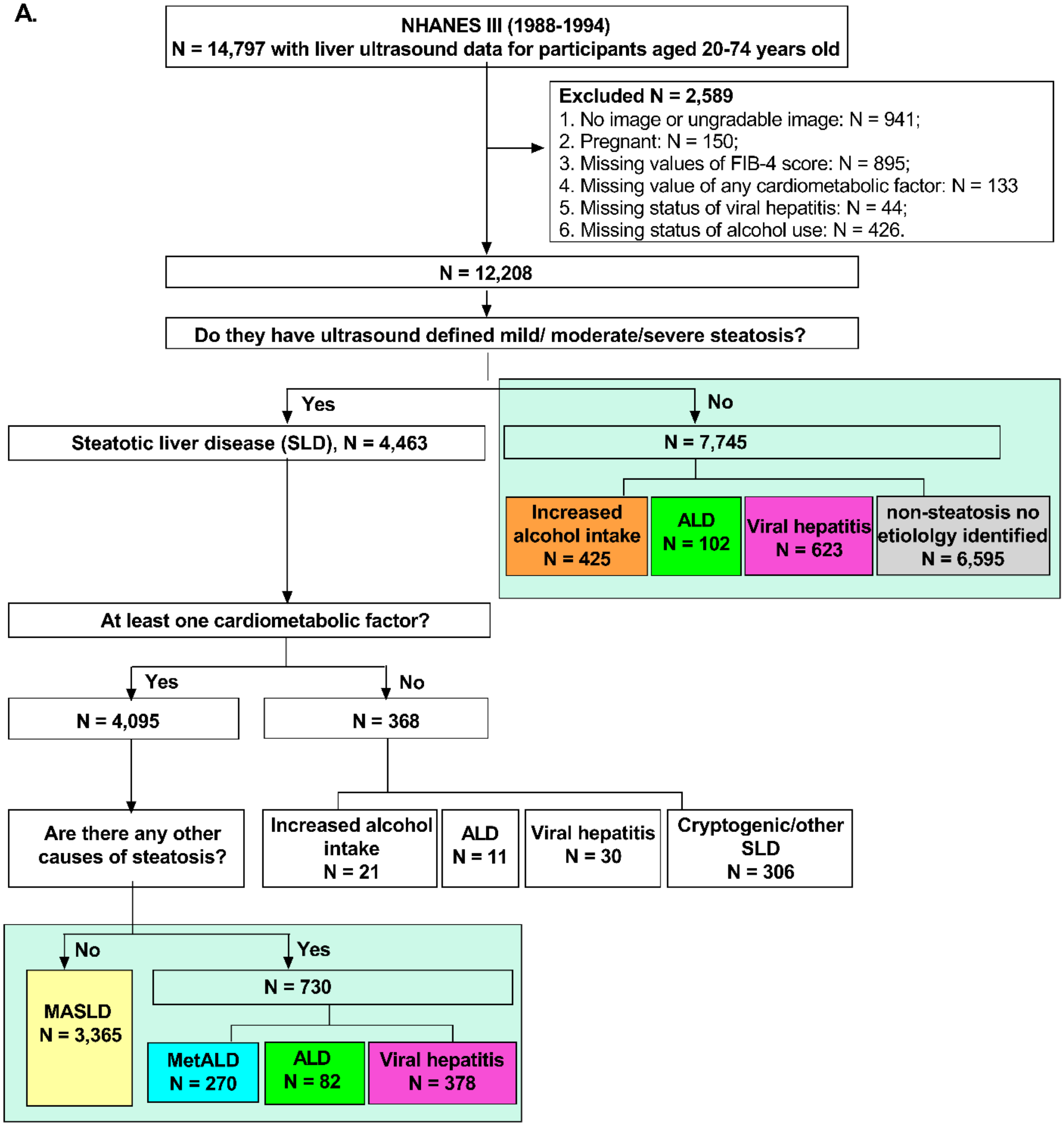
NHANES III (1988-1994)
N = 14,797 with liver ultrasound data for participants aged 20-74 years old
Excluded N = 2,589
1. No image or ungradable image: N = 941;
2. Pregnant: N = 150;
3. Missing values of FIB-4 score: N = 895;
4. Missing value of any cardiometabolic factor: N = 133
5. Missing status of viral hepatitis: N = 44;
6. Missing status of alcohol use: N = 426.
N = 12,208
Do they have ultrasound defined mild/ moderate/severe steatosis?
Yes
No
Steatotic liver disease (SLD), N = 4,463
N = 7,745
Increased alcohol intake N = 425
ALD N = 102
Viral hepatitis N = 623
non-steatosis no etiololgy identified N = 6,595
At least one cardiometabolic factor?
Yes
No
N = 4,095
N = 368
Are there any other causes of steatosis?
Increased alcohol intake N = 21
ALD N = 11
Viral hepatitis N = 30
Cryptogenic/other SLD N = 306
No
Yes
MASLD N = 3,365
N = 730
MetALD N = 270
ALD N = 82
Viral hepatitis N = 378

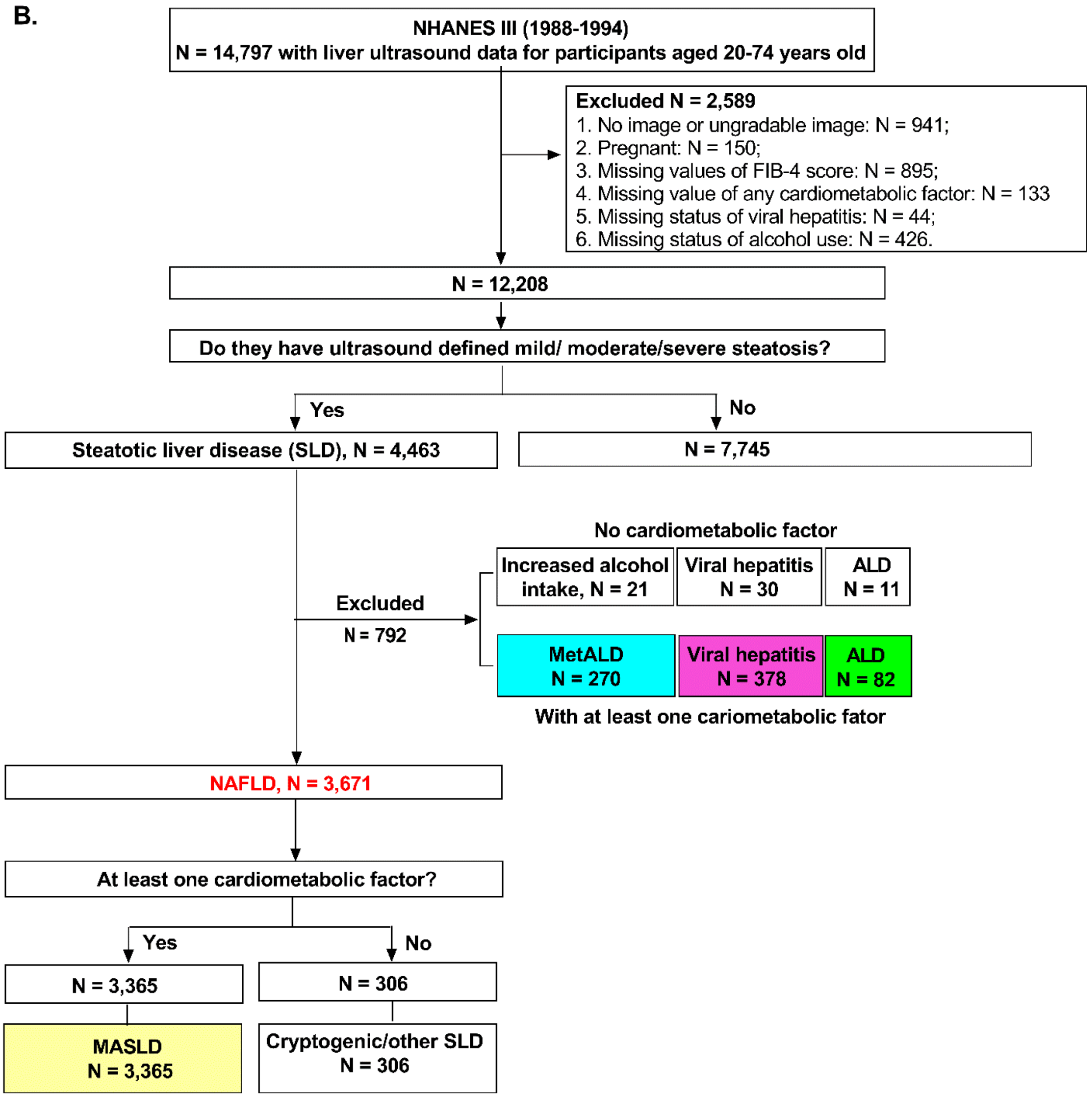

**Figure S3. The new nomenclature's diagnostic scheme applied to adults in the United States with abdominal ultrasound data in NHANES III.** Flowchart of 12,208 adults (20-74 years old) categorized **(A)** according to the proposed new nomenclature; **(B)** according to NAFLD criteria. Abbreviations: Alcohol associated liver disease, ALD; controlled attenuation parameter, CAP; National Health and Nutrition Evaluation Survey, NHANES; Metabolic dysfunction-associated alcohol-associated liver disease, MetALD; Metabolic dysfunction-associated liver disease, MASLD.

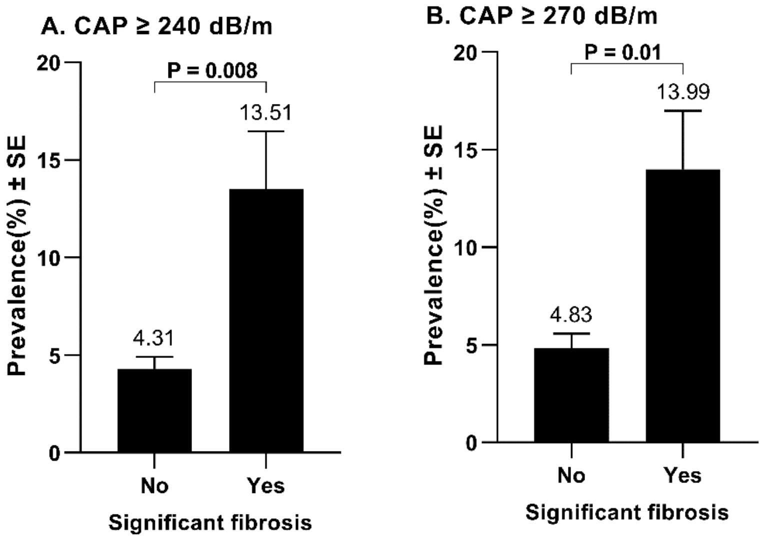

**Figure S4. Age-standardized weighted prevalence of liver disease awareness among adults with steatotic liver disease, stratified by significant fibrosis status.** The age-standardized weighted prevalence of LD awareness was determined in the SLD cohort stratified by significant fibrosis (liver stiffness ≥ 8.6 kPa) status **(A)** Among people with CAP ≥ 240 dB/m, **(B)** Among people with CAP ≥ 270 dB/m**.** Differences between groups were tested by univariate t statistic. Statistical significance was a two-sided P value <0.05. Abbreviations: steatotic liver disease, SLD; standard error, SE.

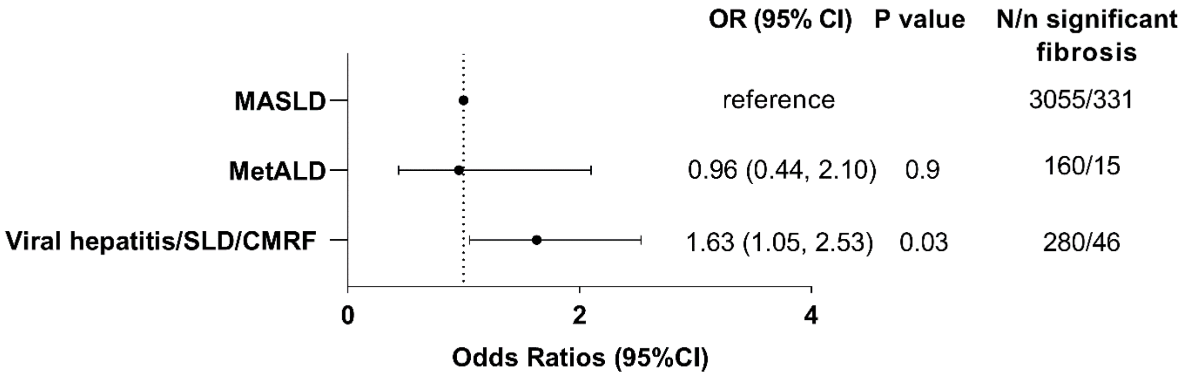

**Figure S5. Unadjusted odds ratios of significant fibrosis among patients with SLD, comparing MASLD, MetALD, and Viral hepatitis/CMRF.** The forest plot shows the crude odds ratios for significant fibrosis for the comparison of MASLD to MetALD and viral hepatitis/SLD/CMRF determined by logistic regression. Significant fibrosis was defined as VCTE-determined liver stiffness ⩾ 8.6 kPa. Abbreviations: cardiometabolic risk factors, CMRF; confidence interval, CI; metabolic dysfunction-associated liver disease, MASLD; metabolic dysfunction-associated alcohol-associated liver disease, MetALD; odds ratio, OR; steatotic liver disease, SLD.

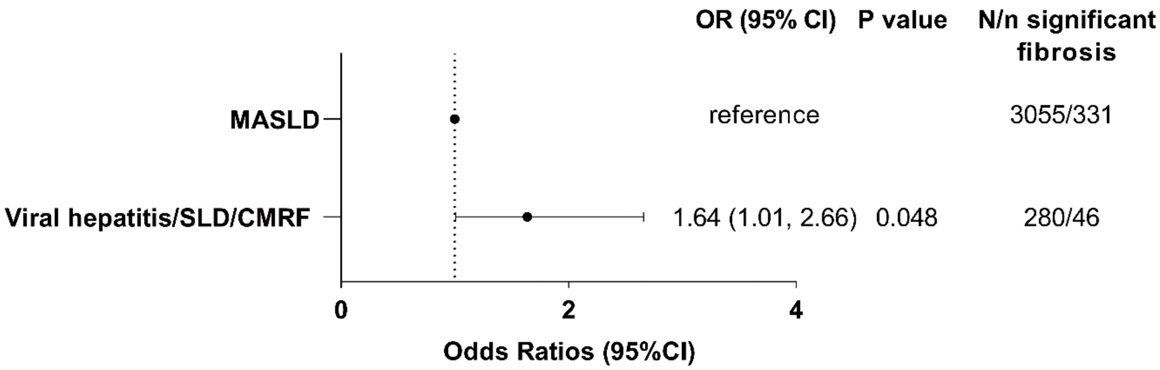

**Figure S6. Adjusted odds ratio of significant fibrosis among patients with SLD, comparing MASLD to viral hepatitis/CMRF, adjusted for age, sex, race/ethnicity.** The forest plot shows the adjusted odds ratio for significant fibrosis for the comparison of MASLD to viral hepatitis/SLD/CMRF determined by logistic regression. Significant fibrosis was defined as VCTE-determined liver stiffness ⩾ 8.6 kPa. Abbreviations: cardiometabolic risk factors, CRMF; confidence interval, CI; metabolic dysfunction-associated liver disease, MASLD; odds ratio, OR; steatotic liver disease, SLD.

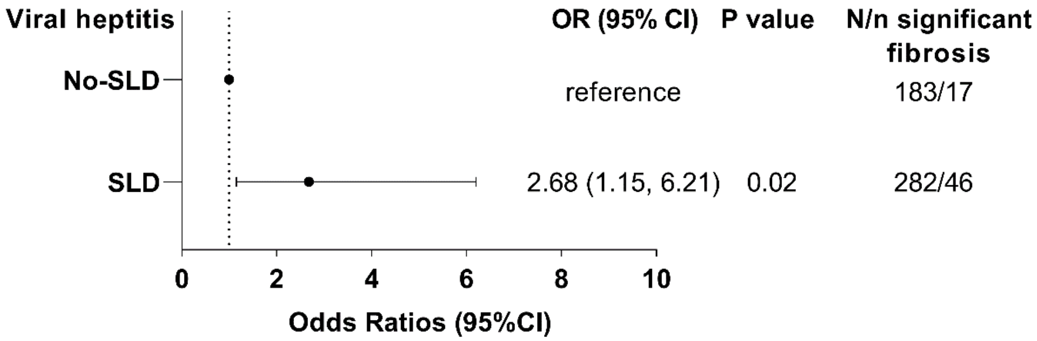

**Figure S7. Adjusted odds ratio significant fibrosis among patients with viral hepatitis, comparing those with and without SLD, adjusted for age.** The forest plot shows the adjusted odds ratio for significant fibrosis for the comparison of viral hepatitis/SLD vs. viral hepatitis/No-SLD based on logistic regression. Significant fibrosis defined by VCTE-determined liver stiffness ⩾ 8.6 kPa. Abbreviations: odds ratio, OR; steatotic liver disease, SLD.

**Table S1. Age-standardized weighted prevalence of elevated adiposity in NHANES III and NHANES 2017-March 2020.**

| BMI/WC criteria | NHANES III (1988-1994) | NHANES 2017-March 2020 |
|---|---|---|
| | Age-standardized weighted % (95%) | |
| **BMI ≥ 25 Kg/m$^2$** | 56.0 (54.4, 57.7) | 74.1 (71.8, 76.3) |
| **WC > 80 cm for women and 94 cm for men** | 60.0 (58.6, 61.4) | 76.8 (73.8, 79.8) |
| **BMI ≥ 25 Kg/m$^2$ or WC > 80 cm for women and 94 cm for men** | 67.0 (65.6, 68.4) | 82.3 (79.7, 85.0) |

Abbreviations: body mass index, BMI; Waist circumference, WC; National Health and Nutrition Evaluation Survey, NHANES

**Table S2. Age-standardized weighted prevalence of liver disease awareness among adults**

| SLD category | Age-standardized weighted % of LD awareness, (95% CI) |
|---|---|
| **SLD (CAP ≥ 240 dB/m)** | 5.3 (1.3, 6.3) |
| **SLD (CAP ≥ 240 dB/m) + LS ≥ 8.0 kPa** | 12.2 (7.0, 17.3) |
| **SLD (CAP ≥ 240 dB/m) + LS ≥ 8.6 kPa** | 13.5 (7.4, 19.6) |
| **SLD (CAP ≥ 270 dB/m)** | 6.1 (4.9, 7.2) |
| **SLD (CAP ≥ 270 dB/m) + LS ≥ 8.0 kPa** | 12.8 (7.4, 18.2) |
| **SLD (CAP ≥ 270 dB/m) + LS ≥ 8.6 kPa** | 14.0 (7.8, 20.2) |

Abbreviations: controlled attenuation parameter, CAP; confidence interval, CI; Steatotic liver disease, SLD; liver stiffness, LS.

**Table S3. Estimated population counts of U.S adolescents with steatotic liver disease and the counts with significant fibrosis**

| SLD category | | Weighted % (95% CI) in total population | Population estimation of each SLD category (million) | Weighted % (95% CI) of LS ≥ 7.4 kPa in each SLD category | Population estimation of LS ≥ 7.4 kPa in each LD category (million) | Weighted % (95% CI) of LS ≥ 8.0 kPa in each SLD category | Population estimation of LS ≥ 8.0 kPa in each LD category (million) |
|---|---|---|---|---|---|---|---|
| **SLD (CAP ≥ 240 dB/m)** | | 30.4 (27.0, 33.9) | 10.3 (9.1, 11.5) | 10.2 (3.0, 17.4) | 1.0 (0.3, 1.8) | 6.8 (1.8, 11.8) | 0.7 (0.2, 1.2) |
| **Sub-categories of SLD (CAP ≥ 240 dB/m)** | **MASLD** | 25.1 (21.8, 28.3) | 8.5 (7.4, 9.6) | 8.9 (3.7, 14.0) | 0.8 (0.3, 1.2) | 6.5 (2.5, 10.6) | 0.6 (0.2, 0.9) |
| | **No CMRF + Cryptogenic/other SLD** | 5.2 (3.4, 6.9) | 1.7 (1.2, 2.3) | 13.5 (0, 32.5) | 0.2 (0, 0.5) | 8.5 (0, 20.0) | 0.1 (0, 0.3) |
| **SLD (CAP ≥ 270 dB/m)** | | 16.4 (13.5, 19.3) | 5.5 (4.6, 6.5) | 14.2 (5.3, 23.0) | 0.8 (0.3, 1.3) | 8.5 (3.5, 13.6) | 0.5 (0.2, 0.7) |
| **Sub-categories of SLD (CAP ≥ 270 dB/m)** | **MASLD** | 15.2 (12.4, 17.9) | 5.1 (4.2, 6.0) | 11.8 (5.4, 18.1) | 0.6 (0.3, 0.9) | 8.1 (3.6, 12.7) | 0.4 (0.2, 0.6) |

Abbreviations: Steatotic liver disease, SLD; cardiometabolic risk factor, CMRF; alcoholic associated liver disease, ALD; Viral hepatitis, VH; metabolic dysfunction associated liver disease, MASLD; Liver stiffness, LS.

**Table S4. Odds ratios of factors associated with significant fibrosis (LS ≥ 8.6 kPa) among adults with MASLD in univariable logistic regression**

| | MASLD | |
|---|---|---|
| **Sample size** | **3,055** | |
| **Significant fibrosis** | **331** | |
| | **OR (95%CI)** | **P value** |
| **Age (unit 10 years)** | 1.08 (0.99, 1.19) | 0.08 |
| **Gender** | | |
| Female | Reference | |
| Male | 1.43 (0.90, 2.28) | 0.13 |
| **Diabetes** | | |
| No | Reference | |
| Yes | 4.31 (3.23, 5.74) | <0.0001 |
| **Hypertension** | | |
| No | Reference | |
| Yes | 2.91 (1.96, 4.33) | <0.0001 |
| **Waist circumference (cm)** | 1.07 (1.06, 1.08) | <0.0001 |
| **BMI (Kg/m$^2$)** | 1.13 (1.10, 1.15) | <0.0001 |
| **Raised triglycerides** | | |
| No | | |
| Yes | 1.27 (0.92, 1.76) | 0.15 |
| **Reduced HDL** | | |
| No | | |
| Yes | 1.16 (0.80, 1.69) | 0.4 |
| **Smoking status** | | |
| Never | Reference | |
| Past/current | 1.06 (0.72, 1.56) | 0.8 |
| **Alcohol use** | | |
| Lifetime abstainers | Reference | |
| Former drinkers | 0.75 (0.40, 1.40) | 0.4 |
| Current drinkers | 0.59 (0.38, 0.93) | 0.02 |
| **Lead** | | |
| Q1-3 | Reference | |
| Q4 | 0.57 (0.41, 0.79) | 0.0007 |
| **Cadmium** | | |
| Q1-3 | Reference | |
| Q4 | 0.61 (0.41, 0.92) | 0.02 |
| **Poverty** | | |
| No | Reference | |
| Yes | 1.20 (0.83, 1.74) | 0.3 |

Note: Multiple imputation was performed in univariable logistic regression models. Variables with P value < 0.1 in the univariable analysis in the in total cohort were included in multivariable models. Abbreviations: body mass index, BMI; High-density lipoprotein cholesterol, HDL; odds ratio, OR; confidence interval, CI; metabolic dysfunction-associated liver disease, MASLD.

**Table S5. Odds ratios of factors associated with significant fibrosis (LS ≥ 8.6 kPa) among adults in the No-SLD No Etiology Identified group in univariable logistic regression**

| | No Etiology Identified | |
|---|---|---|
| **Sample size** | **1,646** | |
| **Significant fibrosis** | **40** | |
| | **OR (95%CI)** | **P value** |
| **Age (unit 10 years)** | 1.17 (0.85, 1.62) | 0.3 |
| **Gender** | | |
| Female | Reference | |
| Male | 2.85 (1.61, 5.07) | 0.0004 |
| **Diabetes** | | |
| No | Reference | |
| Yes | 1.32 (0.52, 3.32) | 0.6 |
| **Hypertension** | | |
| No | Reference | |
| Yes | 1.69 (0.91, 3.14) | 0.1 |
| **Waist circumference (cm)** | 1.05 (1.01, 1.08) | 0.01 |
| **BMI (Kg/m$^2$)** | 1.08 (0.99, 1.19) | 0.08 |
| **Raised triglycerides** | | |
| No | | |
| Yes | 2.91 (1.75, 4.84) | <0.0001 |
| **Reduced HDL** | | |
| No | | |
| Yes | 1.64 (0.97, 2.78) | 0.06 |
| **Smoking status** | | |
| Never | Reference | |
| Past/current | 1.82 (0.44, 7.44) | 0.4 |
| **Alcohol use*** | | |
| Lifetime abstainers | Reference | |
| Former drinkers | - | - |
| Current drinkers | - | - |
| **Lead** | | |
| Q1-3 | Reference | |
| Q4 | 4.84 (2.48, 9.43) | <0.0001 |
| **Cadmium** | | |
| Q1-3 | Reference | |
| Q4 | 1.91 (1.10, 3.32) | 0.02 |
| **Poverty** | | |
| No | Reference | |
| Yes | 0.97 (0.19, 3.93) | 0.9 |

Note: Multiple imputation was performed in univariable logistic regression models. Variables with P value < 0.1 in the univariable analysis in the in total cohort were included in multivariable models. *Only one significant fibrosis case in the reference group of alcohol use, so the estimation was limited by significant fibrosis cases numbers. Abbreviations: body mass index, BMI; High-density lipoprotein cholesterol, HDL; odds ratio, OR; confidence interval, CI; steatotic liver disease, SLD.

**Table S6. Hazard ratio of mortality compared between people with and without steatotic liver disease**

| Mortality type | SLD status | N/ n death | Hazard Ratio (95%) | | | |
|---|---|---|---|---|---|---|
| | | | Unadjusted | Model 1 | Model 2 | Model 3 |
| **All-cause mortality** | No | 7745/2443 | reference | | | |
| | Yes | 4463/1873 | **1.57 (1.46, 1.70)‡** | **1.20 (1.11, 1.31)‡** | 1.07 (0.97, 1.18) | 1.07 (0.98, 1.18) |
| **Heart disease-related mortality** | No | 7745/669 | reference | | | |
| | Yes | 4463/507 | **1.51 (1.29, 1.78)‡** | 1.10 (0.95, 1.28) | 0.89 (0.74, 1.07) | 0.90 (0.76, 1.07) |
| **Cancer-related mortality** | No | 7745/614 | reference | | | |
| | Yes | 4463/426 | **1.40 (1.16, 1.68)†** | 1.11 (0.91, 1.34) | 1.08 (0.86, 1.37) | 1.06 (0.84, 1.34) |

Note: Model 1 adjusted with age, sex, race/ethnicity; Model 2: model 1 plus waist circumference, diabetes and hypertension; Model 3: model 2 plus smoking status and poverty. *P < .05, †P < .001, ‡P < .0001. Abbreviations: steatotic liver disease, SLD.

**Table S7. Hazard ratio of mortality compared between people with no-SLD and those with SLD and at least one CMRF**

| Mortality type | SLD status | N/ n death | Hazard Ratio (95%) | | | |
|---|---|---|---|---|---|---|
| | | | Unadjusted | Model 1 | Model 2 | Model 3 |
| **All-cause mortality** | No | 7745/2443 | reference | | | |
| | SLD/CMRF | 4095/1823 | **1.74 (1.61, 1.89)‡** | **1.21 (1.12, 1.32)‡** | 1.06 (0.97, 1.17) | 1.06 (0.96, 1.16) |
| **Heart disease related mortality** | No | 7745/669 | reference | | | |
| | SLD/CMRF | 4095/498 | **1.70 (1.46, 1.99)‡** | 1.12 (0.97, 1.30) | 0.89 (0.74, 1.07) | 0.89 (0.74, 1.06) |
| **Cancer related mortality** | No | 7745/614 | reference | | | |
| | SLD/CMRF | 4095/413 | **1.52 (1.26, 1.82)‡** | 1.10 (0.91, 1.33) | 1.07 (0.85, 1.35) | 1.07 (0.85, 1.35) |

Note: Model 1 adjusted with age, sex, race/ethnicity; Model 2: model 1 plus waist circumference, diabetes and hypertension; Model 3: model 2 plus smoking status and poverty. *P < .05, †P < .001, ‡P < .0001. Abbreviations: steatotic liver disease, SLD; cardiometabolic risk factor, CMRF.

**Table S8. Hazard ratio of all-cause mortality among people with SLD/CMRF**

| Mortality type | Subtype | N/ n death | Hazard Ratio (95%) | | | |
|---|---|---|---|---|---|---|
| | | | Unadjusted | Model 1 | Model 2 | Model 3 |
| **All-cause mortality** | MASLD | 3365/1441 | reference | | | |
| | MetALD | 270/119 | 1.04 (0.75, 1.45) | 1.20 (0.87, 1.65) | 1.23 (0.90, 1.70) | 1.13 (0.82, 1.56) |
| | ALD | 82/47 | 1.17 (0.64, 2.12) | **1.86 (1.16, 2.99)*** | **2.03 (1.25, 3.28)*** | **1.99 (1.29, 3.06)*** |
| | Viral hepatitis | 378/216 | **1.45 (1.06, 1.98)*** | **1.39 (1.01, 1.92)*** | **1.41 (1.01, 1.96)*** | **1.39 (1.01, 1.91)*** |

Note: Model 1 adjusted with age, sex, race/ethnicity; Model 2: model 1 plus waist circumference, diabetes and hypertension; Model 3: model 2 plus smoking status and poverty. *P < .05, †P < .001, ‡P < .0001. Abbreviations: steatotic liver disease, SLD; cardiometabolic risk factor, CMRF.

**Table S9. Hazard ratio of all-cause mortality compared between people with MetALD and people with No-SLD/increased alcohol intake**

| Mortality type | Liver disease status | N/ n death | Hazard Ratio (95%) | | | |
|---|---|---|---|---|---|---|
| | | | Unadjusted | Model 1 | Model 2 | Model 3 |
| All-cause mortality | No-SLD/Increased alcohol intake | 425/136 | reference | | | |
| | MetALD | 270/119 | **1.72 (1.11, 2.67)*** | 1.22 (0.79, 1.87) | 0.98 (0.62, 1.55) | 0.98 (0.61, 1.58) |

Note: Model 1 adjusted with age, sex, race/ethnicity; Model 2: model 1 plus waist circumference, diabetes and hypertension; Model 3: model 2 plus smoking status and poverty. *P < .05, †P < .001, ‡P < .0001. Abbreviations: steatotic liver disease, SLD; cardiometabolic risk factor, CMRF.

**Table S10. Hazard ratio of all-cause mortality in people with viral hepatitis compared between people with No-SLD and people with SLD/CMRF**

| Mortality type | Viral hepatitis status | N/ n death | Hazard Ratio (95%) | | | |
|---|---|---|---|---|---|---|
| | | | Unadjusted | Model 1 | Model 2 | Model 3 |
| All-cause mortality | No-SLD | 623/268 | reference | | | |
| | SLD/CMRF | 378/216 | **1.59 (1.17, 2.15)*** | 1.30 (0.99, 1.72) | 1.16 (0.85, 1.59) | 1.16 (0.85, 1.58) |

Note: Model 1 adjusted with age, sex, race/ethnicity; Model 2: model 1 plus waist circumference, diabetes and hypertension; Model 3: model 2 plus smoking status and poverty. *P < .05, †P < .001, ‡P < .0001. Abbreviations: steatotic liver disease, SLD; cardiometabolic risk factor, CMRF.

**Table S11. Hazard ratio of all-cause mortality in people with ALD compared between people with No-SLD and with SLD/CMRF**

| Mortality type | ALD status | N/ n death | Hazard Ratio (95%) | | | |
|---|---|---|---|---|---|---|
| | | | Unadjusted | Model 1 | Model 2 | Model 3 |
| All-cause mortality | No-SLD | 102/36 | reference | | | |
| | SLD/CMRF | 82/47 | **1.87 (1.06, 3.28)*** | 1.16 (0.73, 1.86) | NA | NA |

Note: Model 1 adjusted with age, sex, race/ethnicity; Model 2: model 1 plus waist circumference, diabetes and hypertension; Model 3: model 2 plus smoking status and poverty. *P < .05, †P < .001, ‡P < .0001. Abbreviations: steatotic liver disease, SLD; cardiometabolic risk factor, CMRF; not available, NA.

**Table S12. Hazard ratio of all-cause mortality compared between people with No-SLD/No Etiology Identified and people with MASLD**

| Mortality type | Liver disease status | N/ n death | Hazard Ratio (95%) | | | |
|---|---|---|---|---|---|---|
| | | | Unadjusted | Model 1 | Model 2 | Model 3 |
| All-cause mortality | Non-SLD No Etiology Identified | 6595/2003 | reference | | | |
| | MASLD | 3365/1411 | **1.74 (1.60, 1.90)‡** | **1.20 (1.10, 1.31)†** | 1.04 (0.93, 1.16) | 1.04 (0.94, 1.16) |

Note: Model 1 adjusted with age, sex, race/ethnicity; Model 2: model 1 plus waist circumference, diabetes and hypertension; Model 3: model 2 plus smoking status and poverty. *P < .05, †P < .001, ‡P < .0001. Abbreviations: steatotic liver disease, SLD; cardiometabolic risk factor, CMRF.
